# Uptake and Safety Profile of Anti-Amyloid Immunotherapies in Routine Clinical Practice

**DOI:** 10.1101/2025.09.11.25335562

**Authors:** Jay B. Lusk, Kate Vinita Fitch, Kim G Johnson, Andy Liu, Jennifer L. Lund, Laine E. Thomas, Ryan McDevitt, Anqi Zhao, Heather Whitson, Richard O’Brien, Shannon Aymes, Bradley G. Hammill, Brian Mac Grory, Fan Li, Emily C. O’Brien

## Abstract

**Introduction:** Anti**-**amyloid immunotherapies are approved by the United States Food and Drug Administration (FDA) for the treatment of Alzheimer’s disease. The adoption and safety profile of these medications in routine clinical practice have not been described.

**Methods:** We performed a retrospective observational cohort study using nationwide electronic health record data from Epic Cosmos. The principal objective was to describe the baseline characteristics of patients prescribed anti-amyloid immunotherapy in routine clinical practice. Secondarily, we wished to determine whether prescription of anti-amyloid immunotherapy (with or without an acetylcholinesterase inhibitor [AChEI] or memantine) was associated with an increased risk of key safety end points when compared to an AChEI or memantine alone. We used a target trial emulation framework to identify and mitigate sources of bias. The primary end point was time to first nontraumatic intracranial hemorrhage (ICH). Secondary end points included other cardiovascular conditions (ischemic stroke (IS), myocardial infarction (MI), and a composite of ICH, IS, or MI), headache, diarrhea and overall healthcare utilization. Exploratory end points included adverse events linked to other immunotherapies. We used propensity score overlap weighting to balance baseline demographic and clinical characteristics across treatment groups.

**Results:** Between July 1, 2023 and January 1, 2025, 2,616 patients (median age 74.8 years [IQR 69.8-78.8]; 53.9 % female) were prescribed anti-amyloid immunotherapy (with or without AChEI/memantine), and 1,065,192 patients (median age 79.98 [IQR 73.6-85.6], 57.9% female) were prescribed AChEI/memantine alone. In total, 401 patients prescribed anti-amyloid immunotherapy and 274,470 patients prescribed AChEI/memantine were assessed for safety end points. Compared with AChEI/memantine, prescription of anti-amyloid immunotherapy was not associated with increased hazard of ICH after adjustment (owHR 0.73 [95% CI 0.11-5.46]). Anti-amyloid immunotherapy prescription was associated with a higher risk of headache (owHR 2.16 [95% CI 1.12-4.16]) and respiratory infection (owHR 1.57 [95% CI 1.04-2.37]) but was not associated with other immune-related safety endpoints.

**Conclusion:** Anti-amyloid immunotherapy has been principally adopted by patients who are younger and medically healthier than patients receiving AChEI/memantine alone. Prescription of anti-amyloid immunotherapy was not associated with an increased risk of ICH.

## INTRODUCTION

Pivotal trials demonstrated the efficacy of monoclonal antibody therapy targeting amyloid beta (AB) in slowing the clinical progression of Alzheimer’s disease, leading to accelerated approval of aducanumab in June 2021, followed by accelerated approval of lecanemab in January 2023 (with full approval in July 2023), then full approval of donanemab in July 2024.^1–4^ The balance of benefits and harms of these therapies has been a subject of intense debate, particularly due to the risk of amyloid-related imaging abnormalities (ARIA).^5–9^ Although recent studies have estimated low treatment eligibility (approximately 5-8% of patients with mild cognitive impairment or mild dementia),^10–13^, the real-world uptake and safety profile of these medications in the U.S. are not yet known.

It is important to understand the risks of harm associated with this therapy in routine clinical practice.^14–16^ Therefore, the objectives of this study were twofold: 1) to describe the uptake of anti-amyloid monoclonal antibody therapies in routine clinical practice and 2) to determine if the prescription of anti-amyloid immunotherapy was associated with an increased risk of key safety end points compared to an AChEI or memantine.

## METHODS

### Study Design

This was a retrospective observational cohort study which was designed and reported according to a prespecified statistical analysis plan (**Supplemental Material).**

### Data Source

Data used in this study were obtained from Epic Cosmos, a data set created in collaboration with a community of Epic health systems representing more than 300 million patient records from more than 1,748 hospitals and 40,700 clinics from all 50 states, the District of Columbia, Lebanon, and Saudi Arabia. Data available include structured (billing and procedural codes) and semi-structured data (patient demographics, diagnoses, procedures, physical measurements and laboratory measurements) which are harmonized across sites.

### Data Sharing Statement

Data can be obtained from Epic Systems under an approved institutional Data Use Agreement for Epic Cosmos.

### Ethical Approval and Reporting

The study was determined exempt from institutional review board review by the University of North Carolina Chapel Hill institutional review board (review number 459493). The study was reported according to the RECORD and TARGET statements **(Supplemental Material).**

### Study Population

The study population included all patients who received their first prescription for an anti-amyloid monoclonal antibody therapy (lecanemab or donanemab) or an acetylcholinesterase inhibitor (donepezil, rivastigmine, or galantamine) or NMDA receptor antagonist (memantine) between July 1, 2023 and January 1, 2025. The comparator population of patients receiving an acetylcholinesterase inhibitor was selected to allow for alignment of index date and to contextualize the background risk of adverse events unrelated to anti-amyloid immunotherapy. No participants were excluded from the study when evaluating uptake of anti-amyloid therapies. For the safety outcome analysis, a sub-cohort was created by applying a set of exclusion criteria based on a two-year look back period from the initial prescription date. Exclusions included a history of ischemic or hemorrhagic stroke, atrial fibrillation/flutter, cardiac valvular surgery, prescription of warfarin or any non-vitamin K oral anticoagulant, deep venous thrombosis, a hospice/palliative services in the lookback period, or being aged 90 years or older the index date.

### Exposures

The primary exposure was prescription of anti-amyloid monoclonal antibody therapy, either lecanemab or donanemab, between July 1, 2023 and January 1, 2025. The index date was defined as the first prescription date for either an anti-amyloid therapy or an AChEI/memantine. Patients were assigned to one of two groups: those receiving anti-amyloid therapy or a comparator group receiving AChEI/memantine. In cases of combination therapy, patients were assigned to the group for the medication that was prescribed first. A pre-specified sensitivity analysis was performed using a clone-censor-weight approach to account for the fact that many patients were expected to use both therapies.^25^

### End Points

The primary end point was first diagnosis of nontraumatic intracerebral hemorrhage (ICD-10-CM I61.* or ICD-10-CM I62.*). This endpoint was chosen because while Amyloid-Related Imaging Abnormalities (ARIA) do not have a specific ICD-10 code, a severe manifestation of ARIA (such as ARIA-H/hemorrhage) is highly likely to be captured as a non-traumatic intracerebral hemorrhage. Secondary end points included first myocardial infarction, ischemic stroke, and composite of myocardial infarction, ischemic stroke, and intracerebral hemorrhage, diagnosis with diarrheal illness or vomiting, and diagnosis of headache. Additional end points included first emergency department visit or hospitalization as well as count of emergency department visits and hospitalizations over 1 year after first prescription. A negative control (falsification) endpoint of cataract diagnosis was used to assess residual confounding, as it is a common age-related condition that requires health system engagement but is not biologically linked to either treatment.

Exploratory safety end points based on immune-related adverse effects seen in other immunotherapies, including diagnosis of lower or upper respiratory tract infection, myocarditis or pericarditis, acute liver injury, interstitial lung disease, severe skin reactions, hypothyroidism or hyperthyroidism, and hyperosmolar hyperglycemic state or diabetic ketoacidosis. The specific codes for all endpoints are detailed in the **Supplemental Material.**

### Statistical Analysis

Patient characteristics were summarized using counts and percentages for categorical variables and means and standard deviations for continuous variables and standardized mean differences were used to compare characteristics between groups. Propensity score overlap weighting was used to account for measured confounding because it effectively handles extreme propensity scores and ensures covariate balance across groups. ^17,18^ Variables that were collinear or exhibited low variance were omitted as detailed in the **Supplemental Material.** The propensity score model included a set of variables defined using a two-year look-back period, including:

- Demographics: Age, sex, race, and ethnicity
- Social determinants of health: Social vulnerability index, rural urban commuting area (RUCA) classification, marital status, and state/province.
- Physical measurements: Systolic blood pressure, diastolic blood pressure, body mass index, oxygen saturation, and heart rate
- Laboratory measurements: Hemoglobin, hematocrit, white blood cell count, platelet count, aspartate aminotransferase, alanine aminotransferase, serum creatinine, sodium, potassium, glycated hemoglobin, low density lipoprotein cholesterol, high density lipoprotein cholesterol, serum triglycerides
- Prior procedures: Coronary artery bypass grafting, carotid endarterectomy, cardiac ablation, pacemaker placement, and percutaneous coronary intervention, as well as the procedural components of the Claims-based Frailty Index (CFI).^19^
- Prior diagnoses: Individual components of the Elixhauser comorbidity index^20,21^ as well as the individual components of the CFI and common medical conditions affecting patients with cognitive impairment, as detailed in the **Supplemental Material.**
- Current medications: Antihypertensive, diuretic, antithrombotic, lipid lowering, antiglycemic, antidepressant, anxiolytic, antipsychotic, endocrine, respiratory, gastrointestinal, opioid, and antibiotic medications, as detailed in the **Supplemental Material.**
- Prior healthcare utilization: Number of outpatient encounters, emergency department encounters, hospitalizations, and number of visits to neurologists, geriatricians, psychiatrists, primary care doctors, and cardiologists.

Time to event endpoints were analyzed using Cox proportional hazard models, with patients censored at death, end of data availability, or at one year after the index date. For count endpoints, Quasi-Poisson regression was used to account for overdispersion, and the *survey* package was used to implement overlap weights for adjusted models.^22^ Missing data was imputed using random forest-based imputation, given the large number of adjustment variables of different types.^23^ Analyses were performed in Microsoft SQL Server and R with RStudio. The threshold for two-sided tests was set at alpha = 0.05; no adjustment was made for multiple comparisons. E-values were calculated for primary end points to assess the potential impact of unmeasured confounding on study results.^24^

## RESULTS

### Characteristics of Patients Ever Receiving Anti-Amyloid Immunotherapy versus Acetylcholinesterase Inhibitor Therapy Alone

In total, 2,616 patients (median age 74.8, IQR 69.8-78.8, 53.9 % female) received anti-amyloid immunotherapy (with or without AChEI/Memantine), and 1,065,192 patients (median age 79.98, IQR 73.6-85.6, 57.9% female) received AChEI/Memantine alone over the study period. Compared with patients prescribed AChEI/Memantine, those prescribed anti-amyloid immunotherapy were younger (mean age 74 vs. 79, SMD 0.506), less likely to be female (57.9% vs. 53.9%, SMD 0.081), less likely to be Hispanic/Latino (2.2% vs. 6.7%, SMD 0.227), more likely to be White (86.6% vs. 74.1%, SMD 0.317), and less likely to be Asian (1.1% vs. 2.1%, SMD 0.083) or Black (1.8% vs. 9.9%, SMD 0.353),. Those prescribed anti-amyloid therapy were also more likely to be from urban areas (89.1% vs. 84.2%, SMD 0.146), and less likely to be from suburban (6.5% vs. 9.4%, SMD 0.108) or rural areas (3.0% vs. 4.8%, SMD 0.093), more likely to reside in ZIP codes with higher socioeconomic status (mean SVI percentile 46 vs. 56, SMD 0.392), and more likely to be married (80.9% vs. 50.9%, SMD 0.666) instead of divorced, single, or widowed **(Table 1).**

**Table 1.** Characteristics of patients who were ever prescribed anti-amyloid immunotherapy versus those prescribed only acetylcholinesterase inhibitors or memantine.

| Variable | AChEI/Memantine alone<br>(n=1,065,192) | Anti-Amyloid<br>Immunotherapy (n=2,616) | SMD |
| --- | --- | --- | --- |
| **Demographics** |  |  |  |
| Age | 78.55 | 73.92 | 0.506 |
| **Sex** |  |  |  |
| Female, n (%) | 616811 (57.9%) | 1411 (53.9%) | 0.08 |
| Male, n (%) | 448271 (42.1%) | 1205 (46.1%) | 0.08 |
| **Race** |  |  |  |
| American Indian or Alaska Native, n (%) | 2406 (0.2%) | <11 (0%) | 0.067 |
| Asian, n (%) | 22406 (2.1%) | 28 (1.1%) | 0.083 |
| Black or African American, n (%) | 105482 (9.9%) | 46 (1.8%) | 0.353 |
| More than one Race, n (%) | 96274 (9.0%) | 232 (8.9%) | 0.006 |
| Native Hawaiian or Other Pacific Islander, n (%) | 887 (0.1%) | <11 (0%) | 0.018 |
| Other Race, n (%) | 26852 (2.5%) | 17 (0.6%) | 0.15 |
| White, n (%) | 789622 (74.1%) | 2265 (86.6%) | 0.317 |
| **Ethnicity** |  |  |  |
| Hispanic or Latino, n (%) | 71081 (6.7%) | 57 (2.2%) | 0.227 |
| Not Hispanic or Latino, n (%) | 942387 (88.5%) | 2448 (93.6%) | 0.227 |
| **Marital status** |  |  |  |
| Divorced or Legally Separated, n (%) | 98631 (9.3%) | 120 (4.6%) | 0.187 |
| Married, n (%) | 541683 (50.9%) | 2117 (80.9%) | 0.666 |
| Other, n (%) | 32543 (3.1%) | 57 (2.2%) | 0.056 |
| Unmarried, n (%) | 126863 (11.9%) | 104 (4.0%) | 0.299 |
| Widowed, n (%) | 256245 (24.1%) | 210 (8.0%) | 0.452 |
| **Rural-urban commuting area (RUCA) group** |  |  |  |
| Rural, n (%) | 50929 (4.8%) | 79 (3.0%) | 0.093 |
| Suburban, n (%) | 99634 (9.4%) | 170 (6.5%) | 0.108 |
| Urban, n (%) | 896642 (84.2%) | 2332 (89.1%) | 0.146 |
| U.S. Census division |  |  |  |
| East North Central, n (%) | 204904 (19.2%) | 387 (14.8%) | 0.12 |
| East South Central, n (%) | 70929 (6.7%) | 251 (9.6%) | 0.107 |
| Middle Atlantic, n (%) | 132402 (12.4%) | 327 (12.5%) | 0.001 |
| Mountain, n (%) | 49791 (4.7%) | 98 (3.7%) | 0.047 |
| New England, n (%) | 54687 (5.1%) | 233 (8.9%) | 0.148 |
| Pacific, n (%) | 72721 (6.8%) | 110 (4.2%) | 0.116 |
| South Atlantic, n (%) | 268872 (25.2%) | 629 (24.0%) | 0.03 |
| West North Central, n (%) | 63648 (6.0%) | 148 (5.7%) | 0.014 |
| West South Central, n (%) | 135417 (12.7%) | 411 (15.7%) | 0.085 |
| Social Vulnerability Index<br>(percentile, 2020, by ZIP) | 0.56 | 0.46 | 0.392 |
| **Laboratory variables** |  |  |  |
| High-density lipoprotein,<br>mg/dL | 54.56 | 61.08 | 0.367 |
| Low-density lipoprotein,<br>mg/dL | 87.72 | 93.96 | 0.171 |
| Serum Triglycerides, mg/dL | 122.87 | 107.98 | 0.217 |
| Hematocrit (%) | 38.86 | 41.16 | 0.486 |
| Hemoglobin, mg/dL | 12.73 | 13.61 | 0.531 |
| White blood cell count, cells/ul | 8.12 | 6.84 | 0.389 |
| Platelet count, cells/ul | 232.52 | 233.9 | 0.019 |
| Potassium, mEq/L | 4.18 | 4.26 | 0.171 |
| Sodium, mEq/L | 139.25 | 139.64 | 0.115 |
| Serum Creatinine, mg/dL | 1.14 | 0.95 | 0.321 |
| ALT, u/L | 22.59 | 21.35 | 0.046 |
| AST, u/L | 27.33 | 23.93 | 0.114 |
| HbA1c (%) | 6.46 | 6.13 | 0.272 |
| **Medical history** |  |  |  |
| Myocardial infarction, n (%) | 62516 (5.9%) | 52 (2.0%) | 0.201 |
| Congestive heart failure, n (%) | 137393 (12.9%) | 79 (3.0%) | 0.371 |
| Peripheral vascular disease, n (%) | 116374 (10.9%) | 156 (6.0%) | 0.179 |
| Cerebrovascular disease, n (%) | 165915 (15.6%) | 330 (12.6%) | 0.085 |
| Chronic obstructive pulmonary disease (COPD), n (%) | 158510 (14.9%) | 152 (5.8%) | 0.301 |
| Rheumatologic disease, n (%) | 31478 (3.0%) | 70 (2.7%) | 0.017 |
| Peptic ulcer disease, n (%) | 9820 (0.9%) | <11 (0%) | 0.067 |
| Mild liver disease, n (%) | 35398 (3.3%) | 45 (1.7%) | 0.102 |
| Diabetes mellitus (no complications), n (%) | 224053 (21.0%) | 248 (9.5%) | 0.326 |
| Diabetes mellitus with complications, n (%) | 138267 (13.0%) | 138 (5.3%) | 0.27 |
| Paralysis, n (%) | 16615 (1.6%) | <11 (0%) | 0.136 |
| Chronic kidney disease, n (%) | 197198 (18.5%) | 161 (6.2%) | 0.383 |
| Any malignancy (excluding skin), n (%) | 95354 (9.0%) | 188 (7.2%) | 0.065 |
| Moderate to severe liver disease, n (%) | 3417 (0.3%) | <11 (0%) | 0.067 |
| Metastatic solid tumor, n (%) | 19584 (1.8%) | 12 (0.5%) | 0.13 |
| HIV/AIDS, n (%) | 1395 (0.1%) | <11 (0%) | 0.005 |
| Cardiac arrhythmia, n (%) | 285298 (26.8%) | 350 (13.4%) | 0.339 |
| Depression, n (%) | 230530 (21.6%) | 502 (19.2%) | 0.061 |
| Drug abuse, n (%) | 19686 (1.8%) | 17 (0.6%) | 0.108 |
| Psychoses, n (%) | 35725 (3.4%) | 91 (3.5%) | 0.007 |
| Deep venous thrombosis, n (%) | 4537 (0.4%) | <11 (0%) | 0.092 |
| Valvular heart disease, n (%) | 103159 (9.7%) | 166 (6.3%) | 0.123 |
| Atrial fibrillation, n (%) | 158828 (14.9%) | 134 (5.1%) | 0.33 |
| Atrial flutter, n (%) | 17501 (1.6%) | 17 (0.6%) | 0.093 |
| Alcohol use disorder, n (%) | 16430 (1.5%) | 28 (1.1%) | 0.042 |
| Anxiety disorders, n (%) | 192159 (18.0%) | 498 (19.0%) | 0.026 |
| Chronic pain, n (%) | 286131 (26.9%) | 546 (20.9%) | 0.141 |
| Epilepsy, n (%) | 36548 (3.4%) | 28 (1.1%) | 0.16 |
| Fall-related injury, n (%) | 200038 (18.8%) | 287 (11.0%) | 0.221 |
| Malnutrition, n (%) | 92038 (8.6%) | 93 (3.6%) | 0.214 |
| Migraine, n (%) | 28430 (2.7%) | 76 (2.9%) | 0.014 |
| Osteoarthritis, n (%) | 154399 (14.5%) | 318 (12.2%) | 0.069 |
| Peripheral artery disease, n (%) | 111164 (10.4%) | 136 (5.2%) | 0.196 |
| Pneumonia, n (%) | 55140 (5.2%) | 23 (0.9%) | 0.253 |
| Urinary tract infection, n (%) | 113911 (10.7%) | 94 (3.6%) | 0.278 |
| **Procedures** |  |  |  |
| Coronary artery bypass grafting (CABG), n (%) | 434 (0.0%) | <11 (0%) | 0.033 |
| Electrical cardioversion, n (%) | 2066 (0.2%) | <11 (0%) | 0.031 |
| Pacemaker, n (%) | 16563 (1.6%) | 24 (0.9%) | 0.088 |
| Defibrillator, n (%) | 4364 (0.4%) | <11 (0%) | 0.084 |
| Implantable cardiac monitor, n (%) | 4104 (0.4%) | 14 (0.5%) | 0.01 |
| Cardiac valve surgery, n (%) | 1214 (0.1%) | <11 (0%) | 0.056 |
| Percutaneous coronary intervention (stent), n (%) | 3292 (0.3%) | <11 (0%) | 0.028 |
| Peripheral arterial stent, n (%) | 1411 (0.1%) | <11 (0%) | 0.026 |
| **Prescription drug exposures** |  |  |  |
| Angiotensin receptor blocker (ARB), n (%) | 226061 (21.2%) | 510 (19.5%) | 0.043 |
| ACE inhibitor, n (%) | 200477 (18.8%) | 363 (13.9%) | 0.134 |
| Calcium channel blocker, n (%) | 293988 (27.6%) | 460 (17.6%) | 0.241 |
| Beta-blocker, n (%) | 388118 (36.4%) | 598 (22.9%) | 0.301 |
| Potassium-sparing diuretic, n (%) | 51517 (4.8%) | 72 (2.8%) | 0.109 |
| Thiazide diuretic, n (%) | 134339 (12.6%) | 272 (10.4%) | 0.069 |
| Loop diuretic, n (%) | 175578 (16.5%) | 125 (4.8%) | 0.387 |
| Alpha-1 antagonist, n (%) | 21559 (2.0%) | 25 (1.0%) | 0.088 |
| Vasodilator, n (%) | 154544 (14.5%) | 170 (6.5%) | 0.264 |
| Anticoagulant, n (%) | 160447 (15.1%) | 114 (4.4%) | 0.368 |
| Antiplatelets, n (%) | 288794 (27.1%) | 460 (17.6%) | 0.23 |
| Statin, n (%) | 559015 (52.5%) | 1339 (51.2%) | 0.026 |
| Fibrates, n (%) | 20629 (1.9%) | 49 (1.9%) | 0.005 |
| Bile acid sequestrant, n (%) | 15342 (1.4%) | 32 (1.2%) | 0.019 |
| PCSK9 inhibitor, n (%) | 6502 (0.6%) | 49 (1.9%) | 0.114 |
| Ezetimibe, n (%) | 38715 (3.6%) | 144 (5.5%) | 0.09 |
| Icosapent ethyl, n (%) | 5463 (0.5%) | 14 (0.5%) | 0.003 |
| Metformin, n (%) | 123337 (11.6%) | 219 (8.4%) | 0.107 |
| Insulin, n (%) | 160801 (15.1%) | 142 (5.4%) | 0.323 |
| GLP-1 receptor Agonists, n (%) | 41037 (3.9%) | 92 (3.5%) | 0.018 |
| SGLT-2 Inhibitors, n (%) | 52819 (5.0%) | 98 (3.7%) | 0.059 |
| Sulfonylureas, n (%) | 49617 (4.7%) | 58 (2.2%) | 0.134 |
| DDP-IV Inhibitors, n (%) | 31281 (2.9%) | 33 (1.3%) | 0.117 |
| Thiazolidinediones, n (%) | 11733 (1.1%) | 22 (0.8%) | 0.027 |
| SSRI, n (%) | 300844 (28.2%) | 816 (31.2%) | 0.065 |
| SNRI, n (%) | 103454 (9.7%) | 267 (10.2%) | 0.017 |
| Tricyclic antidepressant (TCA), n (%) | 37893 (3.6%) | 69 (2.6%) | 0.053 |
| Monoamine oxidase inhibitor, n (%) | 1713 (0.2%) | <11 (0%) | 0.025 |
| Buspirone, n (%) | 47476 (4.5%) | 121 (4.6%) | 0.008 |
| Benzodiazepines, n (%) | 295224 (27.7%) | 624 (23.9%) | 0.088 |
| Trazodone, n (%) | 122810 (11.5%) | 220 (8.4%) | 0.104 |
| Bupropion, n (%) | 54877 (5.2%) | 196 (7.5%) | 0.096 |
| Mirtazapine, n (%) | 76145 (7.1%) | 76 (2.9%) | 0.195 |
| Gaba Analogs, n (%) | 197340 (18.5%) | 306 (11.7%) | 0.192 |
| Typical Antipsychotics, n (%) | 50468 (4.7%) | 42 (1.6%) | 0.179 |
| Second-generation antipsychotic, n (%) | 168863 (15.9%) | 106 (4.1%) | 0.402 |
| Thyroid hormone replacement, n (%) | 181989 (17.1%) | 399 (15.3%) | 0.05 |
| Bisphosphonate, n (%) | 52874 (5.0%) | 153 (5.8%) | 0.039 |
| Short-acting β-agonist (SABA), n (%) | 231652 (21.7%) | 298 (11.4%) | 0.281 |
| Short-acting muscarinic antagonist (SAMA), n (%) | 127479 (12.0%) | 147 (5.6%) | 0.226 |
| Long-acting β-agonist (LABA), n (%) | 78594 (7.4%) | 108 (4.1%) | 0.14 |
| Long-acting muscarinic antagonist (LAMA), n (%) | 82496 (7.7%) | 149 (5.7%) | 0.082 |
| Inhaled corticosteroid, n (%) | 158163 (14.8%) | 364 (13.9%) | 0.027 |
| Proton pump inhibitor, n (%) | 318146 (29.9%) | 530 (20.3%) | 0.223 |
| H2 receptor blocker, n (%) | 144946 (13.6%) | 286 (10.9%) | 0.082 |
| Opioids, n (%) | 408407 (38.3%) | 768 (29.4%) | 0.191 |
| Penicillin (± beta-lactamase inhibitor), n (%) | 140611 (13.2%) | 313 (12.0%) | 0.037 |
| First Generation Cephalosporins, n (%) | 194752 (18.3%) | 325 (12.4%) | 0.163 |
| 2nd-generation cephalosporin, n (%) | 28700 (2.7%) | 45 (1.7%) | 0.066 |
| 3rd-generation cephalosporin, n (%) | 96408 (9.1%) | 107 (4.1%) | 0.201 |
| Fourth Generation Cephalosporins, n (%) | 12403 (1.2%) | <11 (0%) | 0.125 |
| Fifth Generation Cephalosporins, n (%) | 22 (0.0%) | <11 (0%) | 0.006 |
| Carbapenem, n (%) | 3909 (0.4%) | <11 (0%) | 0.042 |
| Monobactam, n (%) | 412 (0.0%) | <11 (0%) | 0.028 |
| Macrolide, n (%) | 96750 (9.1%) | 221 (8.4%) | 0.022 |
| Fluoroquinolones, n (%) | 129300 (12.1%) | 234 (8.9%) | 0.104 |
| Tetracycline, n (%) | 106518 (10.0%) | 231 (8.8%) | 0.04 |
| Aminoglycoside, n (%) | 19047 (1.8%) | 60 (2.3%) | 0.036 |
| Sulfonamide, n (%) | 6918 (0.6%) | <11 (0%) | 0.05 |
| Lincosamide, n (%) | 32640 (3.1%) | 72 (2.8%) | 0.019 |
| Glycopeptide, n (%) | 52285 (4.9%) | 63 (2.4%) | 0.133 |
| Oxazolidinone, n (%) | 5570 (0.5%) | <11 (0%) | 0.064 |
| Nitroimidazole, n (%) | 44536 (4.2%) | 89 (3.4%) | 0.041 |
| **Healthcare utilization** |  |  |  |
| Hospital admissions | 0.3 | 0.05 | 0.503 |
| Emergency department visits | 0.62 | 0.18 | 0.484 |
| Outpatient face-to-face visits | 4.91 | 6.55 | 0.2 |
| Primary Care visits | 1.15 | 1.14 | 0.002 |
| Neurology visits | 0.4 | 1.15 | 0.581 |
| Psychiatry visits | 0.09 | 0.14 | 0.053 |
| Geriatrics visits | 0.09 | 0.1 | 0.01 |
| Cardiology visits | 0.41 | 0.41 | 0.001 |
| Hospice/palliative care visits | 0.03 | 0 | 0.025 |
| **Vitals** |  |  |  |
| Body Mass Index (kg/m<sup>2</sup>), median value | 27.18 | 26.15 | 0.192 |
| Diastolic blood pressure (mmHg), median value | 72.79 | 73.35 | 0.066 |
| Pulse rate (beats/min), median value | 73.29 | 69.8 | 0.309 |
| Oxygen saturation (SpO<sub>2</sub>, %), median value | 96.8 | 97.36 | 0.32 |
| Systolic blood pressure (mmHg), median value | 132.34 | 129.98 | 0.148 |
| Body Mass Index (kg/m<sup>2</sup>), closest-to-index value | 27.1 | 26.09 | 0.188 |
| Calculated Body Mass Index (kg/m<sup>2</sup>), closest-to-index value | 27.07 | 26.05 | 0.191 |
| Diastolic blood pressure (mmHg), closest-to-index value | 72.81 | 73.03 | 0.022 |
| Pulse rate (beats/min), closest-to-index value | 73.52 | 70.05 | 0.268 |
| Oxygen saturation (SpO<sub>2</sub>, %), closest-to-index value | 96.72 | 97.31 | 0.278 |
| Systolic blood pressure (mmHg), closest-to-index value | 131.32 | 129.04 | 0.121 |
| Weight (lbs), closest-to-index value | 166.14 | 165.39 | 0.019 |
| Respiratory rate (breaths/min), closest-to-index value | 17.13 | 16.79 | 0.158 |
| Temperature (°F), closest-to-index value | 97.79 | 97.78 | 0.011 |
| Systolic blood pressure (mmHg), median $\geq 130$, n (%) | 352291 (33.1%) | 996 (38.1%) | 0.127 |
| Systolic blood pressure (mmHg), median $\geq 140$, n (%) | 194209 (18.2%) | 491 (18.8%) | 0.142 |
| Systolic blood pressure (mmHg), max $\geq 160$, n (%) | 216238 (20.3%) | 408 (15.6%) | 0.312 |
| Systolic blood pressure (mmHg), max $\geq 180$, n (%) | 90257 (8.5%) | 107 (4.1%) | 0.3 |
| Systolic blood pressure (mmHg), min $\leq 89$, n (%) | 38126 (3.6%) | 39 (1.5%) | 0.207 |
| Oxygen saturation (SpO<sub>2</sub>, %), min $\leq 91$, n (%) | 90064 (8.5%) | 71 (2.7%) | 0.395 |
| Oxygen saturation (SpO<sub>2</sub>, %), min $\leq 89$, n (%) | 40882 (3.8%) | 25 (1.0%) | 0.282 |
| Diastolic blood pressure (mmHg), median $\geq 80$, n (%) | 147754 (13.9%) | 470 (18.0%) | 0.004 |
| Diastolic blood pressure<br>(mmHg), median $\geq 90$, n (%) | 23213 (2.2%) | 65 (2.5%) | 0.025 |
| Diastolic blood pressure<br>(mmHg), max $\geq 100$, n (%) | 81356 (7.6%) | 96 (3.7%) | 0.284 |
| Diastolic blood pressure<br>(mmHg), min $\leq 59$, n (%) | 216757 (20.3%) | 486 (18.6%) | 0.222 |

Patients prescribed anti-amyloid immunotherapy were generally healthier, with lower prevalence of most comorbidities, including heart failure (3.0% vs. 12.9%, SMD 0.371), chronic obstructive pulmonary disease (5.8% vs. 14.9%, SMD 0.301), or atrial fibrillation (5.1% vs. 14.9%, SMD 0.330). Patients prescribed anti-amyloid immunotherapy also had baseline vital measurements consistent with better overall health status, such as lower blood pressure, lower pulse rates, and lower prevalence of documented hypoxia. In terms of medications, patients prescribed anti-amyloid immunotherapy had lower rates of prescription of many chronic disease medications, notably anticoagulants (4.4% vs. 15.1%, SMD 0.368) and antiplatelets (17.6% vs. 27.1%, SMD 0.230), insulin (5.4% vs. 15.1%, SMD 0.323), atypical antipsychotics (4.1% vs. 15.9%, SMD 0.402), and mirtazapine (2.9% vs. 7.1%, SMD 0.195). Patients prescribed anti-amyloid immunotherapy had lower rates of preceding ED visits and hospitalizations and higher number of outpatient visits, especially neurology visits. Characteristics of patients receiving only anti-amyloid immunotherapy versus only AChEI/Memantine are shown in **Supplemental Table 1.**

### Characteristics of patients included in safety outcomes sub-cohort

After the application of safety analysis inclusion and exclusion criteria (**Figure 1),** 401 patients were included in the anti-amyloid immunotherapy group and 274,470 patients were included in the acetylcholinesterase inhibitor group. Patient characteristics before propensity score overlap weighting are shown in **Table 2**; characteristics after overlap weighting are shown in **Supplemental Table 2.**

**Figure 1.**
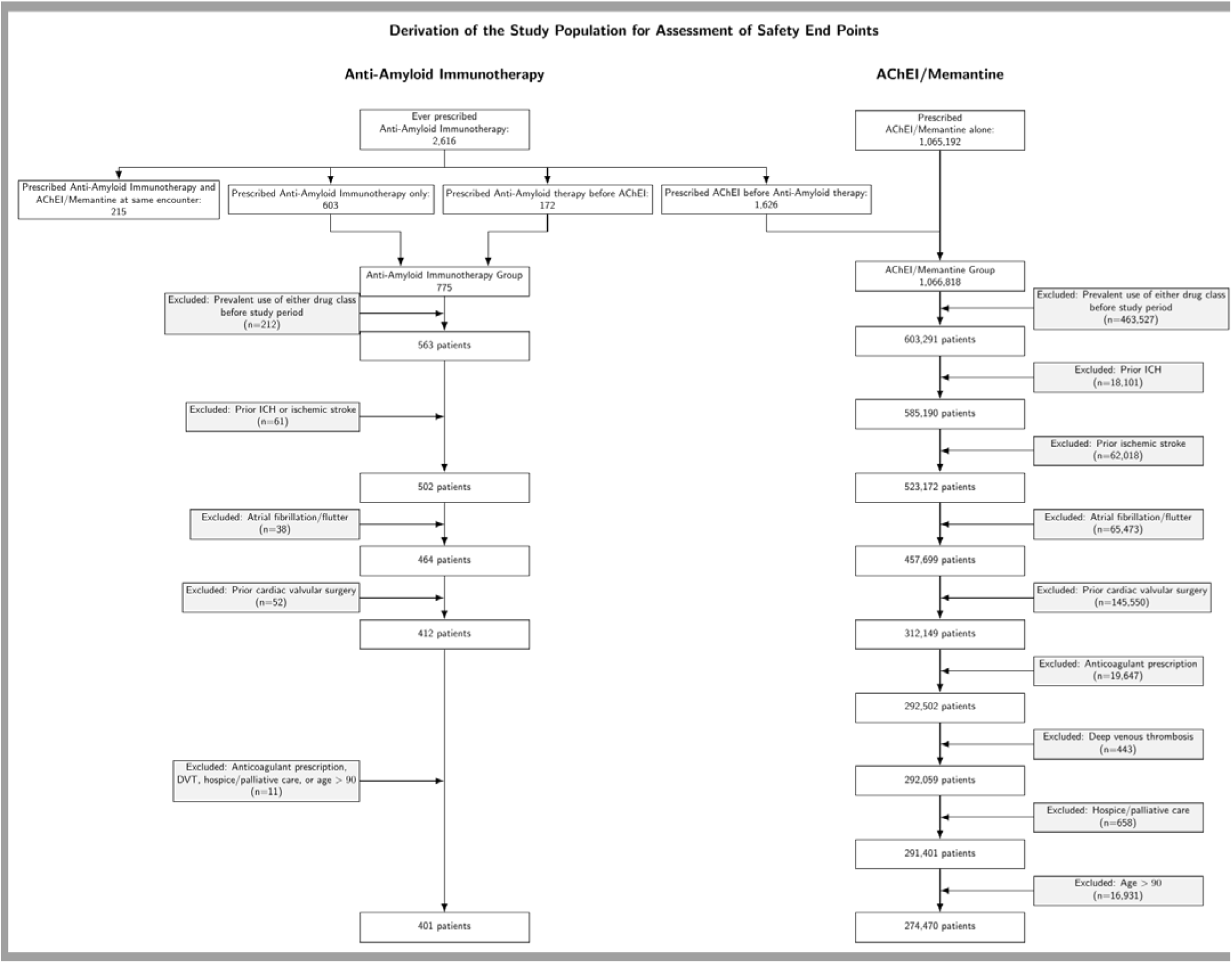
Derivation of the patient population used for safety analysis.

**Table 2.** Characteristics of patients in the safety evaluation sub-cohort before overlap weighting.

| Variable | AChEi/Memantine<br>(N=274,470) | Anti-amyloid immunotherapy<br>(N=401) | SMD |
| --- | --- | --- | --- |
| **Demographics** |  |  |  |
| Age | 75.07 | 73.51 | 0.161 |
| Sex |  |  |  |
| Female, n (%) | 163307 (59.5%) | 223 (55.6%) | 0.079 |
| Male, n (%) | 111132 (40.5%) | 178 (44.4%) | 0.079 |
| Race |  |  |  |
| American Indian or Alaska Native, n (%) | 715 (0.3%) | <11 (0%) | 0.072 |
| Asian, n (%) | 6757 (2.5%) | <11 (0%) | 0.112 |
| Black or African American, n (%) | 24889 (9.1%) | <11 (0%) | 0.328 |
| More than one Race, n (%) | 26993 (9.8%) | 45 (11.2%) | 0.045 |
| Native Hawaiian or Other Pacific Islander, n (%) | 269 (0.1%) | <11 (0%) | 0.036 |
| Other Race, n (%) | 7782 (2.8%) | <11 (0%) | 0.158 |
| White, n (%) | 200701 (73.1%) | 339 (84.5%) | 0.282 |
| Ethnicity |  |  |  |
| Hispanic or Latino, n (%) | 21212 (7.7%) | <11 (0%) | 0.3 |
| Not Hispanic or Latino, n (%) | 253258 (92.3%) | 395 (98.5%) | 0.3 |
| Marital status |  |  |  |
| Divorced or Legally Separated, n (%) | 26271 (9.6%) | 17 (4.2%) | 0.211 |
| Married, n (%) | 151296 (55.1%) | 309 (77.1%) | 0.476 |
| Other, n (%) | 10589 (3.9%) | <11 (0%) | 0.111 |
| Unmarried, n (%) | 36587 (13.3%) | 31 (7.7%) | 0.183 |
| Widowed, n (%) | 49727 (18.1%) | 36 (9.0%) | 0.269 |
| RUCA group |  |  |  |
| Rural, n (%) | 13538 (4.9%) | <11 (0%) | 0.178 |
| Suburban, n (%) | 27335 (10.0%) | 27 (6.7%) | 0.117 |
| Urban, n (%) | 233597 (85.1%) | 367 (91.5%) | 0.201 |
| U.S. Census division |  |  |  |
| East North Central, n (%) | 50079 (18.2%) | 52 (13.0%) | 0.146 |
| East South Central, n (%) | 18012 (6.6%) | 22 (5.5%) | 0.045 |
| Middle Atlantic, n (%) | 32884 (12.0%) | 63 (15.7%) | 0.108 |
| Mountain, n (%) | 13356 (4.9%) | 12 (3.0%) | 0.097 |
| New England, n (%) | 13987 (5.1%) | 31 (7.7%) | 0.108 |
| Pacific, n (%) | 19860 (7.2%) | 21 (5.2%) | 0.083 |
| South Atlantic, n (%) | 72603 (26.5%) | 122 (30.4%) | 0.088 |
| West North Central, n (%) | 16706 (6.1%) | 17 (4.2%) | 0.084 |
| West South Central, n (%) | 36983 (13.5%) | 61 (15.2%) | 0.05 |
| Social Vulnerability Index<br>(percentile, 2020, by ZIP) | 0.57 | 0.45 | 0.438 |
| **Laboratory variables** |  |  |  |
| High-density lipoprotein,<br>mg/dL | 57.52 | 58.59 | 0.061 |
| Low-density lipoprotein,<br>mg/dL | 98.92 | 97.7 | 0.033 |
| Serum Triglycerides, mg/dL | 119.61 | 113.18 | 0.094 |
| Hematocrit (%) | 40.88 | 42.24 | 0.297 |
| Hemoglobin, mg/dL | 13.44 | 13.95 | 0.325 |
| White blood cell count, cells/ul | 7.12 | 6.76 | 0.134 |
| Platelet count, cells/ul | 238.51 | 236.75 | 0.024 |
| Potassium, mEq/L | 4.24 | 4.29 | 0.093 |
| Sodium, mEq/L | 139.6 | 139.73 | 0.046 |
| Serum Creatinine, mg/dL | 0.99 | 0.93 | 0.147 |
| ALT, u/L | 20.15 | 20.37 | 0.013 |
| AST, u/L | 23.61 | 22.8 | 0.046 |
| HbA1c (%) | 5.94 | 5.8 | 0.152 |
| **Medical history** |  |  |  |
| Myocardial infarction, n (%) | 7890 (2.9%) | <11 (0%) | 0.057 |
| Congestive heart failure, n (%) | 14892 (5.4%) | <11 (0%) | 0.182 |
| Peripheral vascular disease, n (%) | 21388 (7.8%) | 31 (7.7%) | 0.002 |
| Cerebrovascular disease, n (%) | 9965 (3.6%) | 14 (3.5%) | 0.008 |
| Chronic obstructive pulmonary disease (COPD), n (%) | 30178 (11.0%) | 23 (5.7%) | 0.191 |
| Rheumatologic disease, n (%) | 7630 (2.8%) | 11 (2.7%) | 0.002 |
| Peptic ulcer disease, n (%) | 1696 (0.6%) | <11 (0%) | 0.112 |
| Mild liver disease, n (%) | 8373 (3.1%) | 13 (3.2%) | 0.011 |
| Diabetes mellitus (no complications), n (%) | 47662 (17.4%) | 48 (12.0%) | 0.153 |
| Diabetes mellitus with complications, n (%) | 25932 (9.4%) | 24 (6.0%) | 0.13 |
| Paralysis, n (%) | 1335 (0.5%) | <11 (0%) | 0.002 |
| Chronic kidney disease, n (%) | 32137 (11.7%) | 22 (5.5%) | 0.223 |
| Any malignancy (excluding skin), n (%) | 25713 (9.4%) | 26 (6.5%) | 0.107 |
| Moderate to severe liver disease, n (%) | 796 (0.3%) | <11 (0%) | 0.076 |
| Metastatic solid tumor, n (%) | 5775 (2.1%) | <11 (0%) | 0.115 |
| HIV/AIDS, n (%) | 380 (0.1%) | <11 (0%) | 0.025 |
| Cardiac arrhythmia, n (%) | 28102 (10.2%) | 34 (8.5%) | 0.06 |
| Depression, n (%) | 52803 (19.2%) | 64 (16.0%) | 0.086 |
| Drug abuse, n (%) | 4938 (1.8%) | <11 (0%) | 0.094 |
| Psychoses, n (%) | 8255 (3.0%) | 18 (4.5%) | 0.078 |
| Valvular heart disease n (%) | 14843 (5.4%) | 25 (6.2%) | 0.035 |
| Alcohol use disorder n (%) | 3838 (1.4%) | <11 (0%) | 0.013 |
| Alzheimer's Disease diagnosis, n (%) | 41943 (15.3%) | 225 (56.1%) | 0.942 |
| Anxiety Disorders, n (%) | 48287 (17.6%) | 72 (18.0%) | 0.009 |
| Chronic pain, n (%) | 73170 (26.7%) | 101 (25.2%) | 0.034 |
| Epilepsy, n (%) | 5867 (2.1%) | <11 (0%) | 0.144 |
| Fall-related injury, n (%) | 34678 (12.6%) | 47 (11.7%) | 0.028 |
| Malnutrition, n (%) | 17276 (6.3%) | 15 (3.7%) | 0.117 |
| Migraine, n (%) | 10002 (3.6%) | 17 (4.2%) | 0.031 |
| Osteoarthritis, n (%) | 35600 (13.0%) | 48 (12.0%) | 0.03 |
| Peripheral artery disease, n (%) | 20802 (7.6%) | 24 (6.0%) | 0.063 |
| Pneumonia, n (%) | 6034 (2.2%) | <11 (0%) | 0.148 |
| Urinary Tract Infection, n (%) | 16208 (5.9%) | 18 (4.5%) | 0.064 |
| **Procedures** |  |  |  |
| Coronary artery bypass grafting (CABG), n (%) | 63 (0.0%) | <11 (0%) | 0.021 |
| Electrical cardioversion, n (%) | 20 (0.0%) | <11 (0%) | 0.012 |
| Pacemaker, n (%) | 1740 (0.6%) | <11 (0%) | 0.014 |
| Defibrillator, n (%) | 583 (0.2%) | <11 (0%) | 0.008 |
| Implantable cardiac monitor, n (%) | 319 (0.1%) | <11 (0%) | 0.096 |
| Percutaneous coronary intervention (stent), n (%) | 855 (0.3%) | <11 (0%) | 0.012 |
| Peripheral arterial stent, n (%) | 287 (0.1%) | <11 (0%) | 0.046 |
| **Prescription drug exposures** |  |  |  |
| Angiotensin receptor blocker (ARB), n (%) | 54883 (20.0%) | 81 (20.2%) | 0.005 |
| ACE inhibitor, n (%) | 47776 (17.4%) | 57 (14.2%) | 0.088 |
| Calcium channel blocker, n (%) | 62135 (22.6%) | 66 (16.5%) | 0.156 |
| Beta-blocker, n (%) | 76941 (28.0%) | 94 (23.4%) | 0.105 |
| Potassium-sparing diuretic, n (%) | 10147 (3.7%) | 14 (3.5%) | 0.011 |
| Thiazide diuretic, n (%) | 36131 (13.2%) | 36 (9.0%) | 0.134 |
| Loop diuretic, n (%) | 25951 (9.5%) | 16 (4.0%) | 0.22 |
| Alpha-1 antagonist, n (%) | 4561 (1.7%) | <11 (0%) | 0.025 |
| Vasodilator, n (%) | 24795 (9.0%) | 28 (7.0%) | 0.076 |
| Antiplatelets, n (%) | 52653 (19.2%) | 71 (17.7%) | 0.038 |
| Statin, n (%) | 129074 (47.0%) | 207 (51.6%) | 0.092 |
| Fibrates, n (%) | 5114 (1.9%) | <11 (0%) | 0.027 |
| Bile acid sequestrant, n (%) | 3160 (1.2%) | <11 (0%) | 0.015 |
| PCSK9 inhibitor, n (%) | 1894 (0.7%) | 20 (5.0%) | 0.261 |
| Ezetimibe, n (%) | 10097 (3.7%) | 33 (8.2%) | 0.193 |
| Icosapent ethyl, n (%) | 1461 (0.5%) | <11 (0%) | 0.076 |
| Metformin, n (%) | 33115 (12.1%) | 45 (11.2%) | 0.026 |
| Insulin, n (%) | 29348 (10.7%) | 29 (7.2%) | 0.121 |
| GLP-1 Agonists, n (%) | 12596 (4.6%) | 20 (5.0%) | 0.019 |
| SGLT-2 Inhibitors, n (%) | 13374 (4.9%) | 17 (4.2%) | 0.03 |
| Sulfonylureas, n (%) | 12316 (4.5%) | <11 (0%) | 0.109 |
| DDP-IV Inhibitors, n (%) | 7226 (2.6%) | <11 (0%) | 0.025 |
| Thiazolidinediones, n (%) | 3134 (1.1%) | <11 (0%) | 0.031 |
| SSRI, n (%) | 69853 (25.5%) | 110 (27.4%) | 0.045 |
| SNRI, n (%) | 27070 (9.9%) | 46 (11.5%) | 0.052 |
| Tricyclic antidepressant (TCA), n (%) | 11050 (4.0%) | <11 (0%) | 0.119 |
| Monoamine oxidase inhibitor, n (%) | 502 (0.2%) | <11 (0%) | 0.061 |
| Buspirone, n (%) | 11592 (4.2%) | 22 (5.5%) | 0.059 |
| Benzodiazepines, n (%) | 68013 (24.8%) | 103 (25.7%) | 0.021 |
| Trazodone, n (%) | 27307 (9.9%) | 32 (8.0%) | 0.069 |
| Bupropion, n (%) | 15582 (5.7%) | 32 (8.0%) | 0.091 |
| Mirtazapine, n (%) | 14219 (5.2%) | <11 (0%) | 0.172 |
| Gaba Analogs, n (%) | 48822 (17.8%) | 53 (13.2%) | 0.127 |
| Typical Antipsychotics, n (%) | 8140 (3.0%) | <11 (0%) | 0.045 |
| Atypical antipsychotic, n (%) | 31555 (11.5%) | 13 (3.2%) | 0.32 |
| Thyroid hormone replacement, n (%) | 41858 (15.3%) | 54 (13.5%) | 0.051 |
| Bisphosphonate, n (%) | 14252 (5.2%) | 20 (5.0%) | 0.009 |
| Short-acting β-agonist (SABA), n (%) | 47647 (17.4%) | 50 (12.5%) | 0.138 |
| Short-acting muscarinic antagonist (SAMA), n (%) | 20869 (7.6%) | 17 (4.2%) | 0.143 |
| Long-acting β-agonist (LABA), n (%) | 18326 (6.7%) | 15 (3.7%) | 0.132 |
| Long-acting muscarinic antagonist (LAMA), n (%) | 18121 (6.6%) | 15 (3.7%) | 0.129 |
| Inhaled corticosteroid, n (%) | 38232 (13.9%) | 52 (13.0%) | 0.028 |
| Proton pump inhibitor, n (%) | 71832 (26.2%) | 92 (22.9%) | 0.075 |
| H2 receptor blocker, n (%) | 32068 (11.7%) | 52 (13.0%) | 0.039 |
| Opioids, n (%) | 87592 (31.9%) | 122 (30.4%) | 0.032 |
| Penicillin (± beta-lactamase inhibitor), n (%) | 32050 (11.7%) | 56 (14.0%) | 0.068 |
| First Generation Cephalosporins, n (%) | 40233 (14.7%) | 47 (11.7%) | 0.087 |
| 2nd-generation cephalosporin, n (%) | 5023 (1.8%) | <11 (0%) | 0.012 |
| 3rd-generation cephalosporin, n (%) | 16031 (5.8%) | 20 (5.0%) | 0.038 |
| Fourth Generation Cephalosporins, n (%) | 1449 (0.5%) | <11 (0%) | 0.045 |
| Fifth Generation Cephalosporins, n (%) | <11 (0%) | <11 (0%) | 0.005 |
| Carbapenem, n (%) | 535 (0.2%) | <11 (0%) | 0.012 |
| Monobactam, n (%) | 68 (0.0%) | <11 (0%) | 0.022 |
| Macrolide, n (%) | 22925 (8.4%) | 42 (10.5%) | 0.073 |
| Fluoroquinolones, n (%) | 28317 (10.3%) | 40 (10.0%) | 0.011 |
| Tetracycline, n (%) | 23623 (8.6%) | 43 (10.7%) | 0.072 |
| Aminoglycoside, n (%) | 4340 (1.6%) | <11 (0%) | 0.048 |
| Sulfonamide, n (%) | 1466 (0.5%) | <11 (0%) | 0.005 |
| Lincosamide, n (%) | 7637 (2.8%) | 16 (4.0%) | 0.067 |
| Glycopeptide, n (%) | 8721 (3.2%) | <11 (0%) | 0.132 |
| Oxazolidinone, n (%) | 750 (0.3%) | <11 (0%) | 0.036 |
| Nitroimidazole, n (%) | 9334 (3.4%) | 16 (4.0%) | 0.031 |
| **Healthcare utilization** |  |  |  |
| Hospital admissions, mean | 0.12 | 0.03 | 0.259 |
| ED Visits, mean | 0.34 | 0.18 | 0.214 |
| Outpatient face-to-face visits, mean | 4.96 | 8.03 | 0.375 |
| Primary care visits, mean | 1.16 | 1.18 | 0.012 |
| Neurology visits, mean | 0.5 | 1.1 | 0.435 |
| Psychiatry visits, mean | 0.12 | 0.12 | 0.002 |
| Geriatrics visits, mean | 0.06 | 0.08 | 0.017 |
| Cardiology visits, mean | 0.27 | 0.57 | 0.176 |
| **Vitals** |  |  |  |
| Body Mass Index (kg/m<sup>2</sup>), median value | 26.85 | 26.5 | 0.063 |
| Diastolic blood pressure (mmHg), median value | 72.89 | 73.9 | 0.122 |
| Pulse rate (beats/min), median value | 72.45 | 70.98 | 0.128 |
| Oxygen saturation (SpO<sub>2</sub>, %), median value | 96.51 | 97.08 | 0.232 |
| Systolic blood pressure (mmHg), median value | 133.47 | 129.88 | 0.235 |
| Body Mass Index (kg/m<sup>2</sup>), closest-to-index value | 26.79 | 26.48 | 0.055 |
| Diastolic blood pressure (mmHg), closest-to-index value | 72.78 | 73.41 | 0.062 |
| Pulse rate (beats/min), closest-to-index value | 72.57 | 71.42 | 0.088 |
| Oxygen saturation (SpO<sub>2</sub>, %), closest-to-index value | 96.46 | 97.01 | 0.213 |
| Systolic blood pressure (mmHg), closest-to-index value | 132.75 | 129.07 | 0.199 |
| Systolic blood pressure (mmHg), median $\geq 130$, n (%) | 147265 (53.7%) | 184 (45.9%) | 0.156 |
| Systolic blood pressure (mmHg), median $\geq 140$, n (%) | 84782 (30.9%) | 83 (20.7%) | 0.235 |
| Systolic blood pressure (mmHg), max $\geq 160$, n (%) | 85894 (31.3%) | 75 (18.7%) | 0.294 |
| Systolic blood pressure (mmHg), max $\geq 180$, n (%) | 24415 (8.9%) | 21 (5.2%) | 0.143 |
| Systolic blood pressure (mmHg), min $\leq 89$, n (%) | 6773 (2.5%) | <11 (0%) | 0.137 |
| Oxygen saturation (SpO<sub>2</sub>, %), min $\leq 91$, n (%) | 50846 (18.5%) | 30 (7.5%) | 0.333 |
| Oxygen saturation (SpO<sub>2</sub>, %), min $\leq 89$, n (%) | 7542 (2.7%) | <11 (0%) | 0.179 |
| Diastolic blood pressure (mmHg), median $\geq 80$, n (%) | 52925 (19.3%) | 102 (25.4%) | 0.148 |
| Diastolic blood pressure (mmHg), median $\geq 90$, n (%) | 6951 (2.5%) | <11 (0%) | 0.002 |
| Diastolic blood pressure (mmHg), max $\geq 100$, n (%) | 15984 (5.8%) | 12 (3.0%) | 0.138 |
| Diastolic blood pressure (mmHg), min $\leq 59$, n (%) | 72613 (26.5%) | 84 (20.9%) | 0.13 |

### Primary End Point

Crude and adjusted rates of ICH are shown in **Figure 2**. Crude and adjusted survival curves are shown in **Supplemental Figure 1.** The crude rate of intracranial hemorrhage was 3.82 per 1,000 person-years in the AChEI/memantine group and 2.95 in the anti-amyloid immunotherapy group (HR 0.77, 95% CI 0.11-5.46). The overlap-weighting adjusted rate of intracranial hemorrhage was 4.00 per 1,000 person-years in the AChEI/memantine group and 2.97 in the anti-amyloid immunotherapy group (owHR 0.73, 95% CI 0.10-5.12).

**Figure 2.**
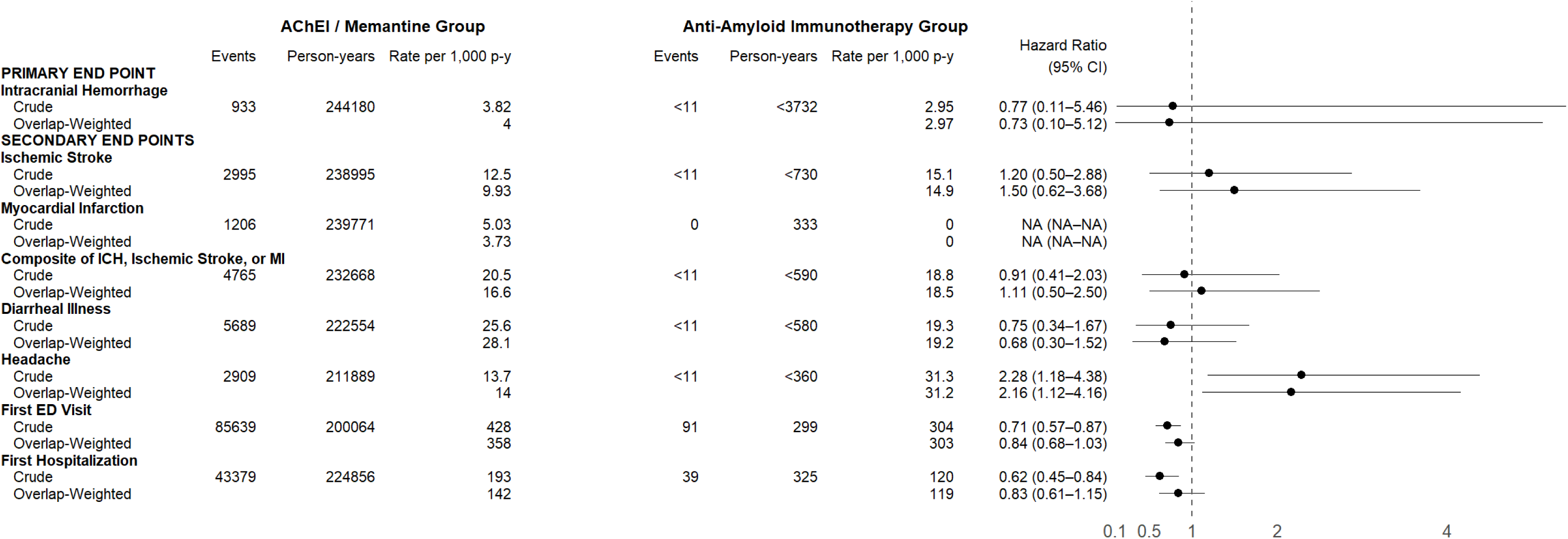
Crude and adjusted rates and hazard ratios for the primary and secondary study end points.

### Secondary End Points

There was no association between anti-amyloid immunotherapy prescription and rates of ischemic stroke, myocardial infarction, or the composite of ICH, ischemic stroke, or MI before or after overlap weighting **(Figure 2).** Crude and adjusted survival curves are shown in **Supplemental Figure 1.** Furthermore, there was no association between anti-amyloid immunotherapy and diagnosis of diarrheal illness before or after overlap weighting (**Figure 2)**.

There was an increased rate of headache (owHR 2.28, ([95% CI 1.18-4.38]; adjusted rate 14.0 per 1,000 p-y vs. 31.2 per 1,000 p-y, adjusted rate difference 17.2 per 1,000 p-y, NNH of 58) after overlap weighting (**Figure 2).** Anti-amyloid immunotherapy prescription was associated with lower hazard of emergency department visit (HR 0.71, 95% CI 0.57-0.87) and hospitalization (HR 0.62, 95% CI 0.45-0.84) before adjustment, but after adjustment these differences were attenuated (owHR 0.84, 95% CI 0.68-1.03 and owHR 0.83, 95% CI 0.61-1.15 respectively).

Patterns of healthcare utilization are shown in **Supplemental Table 3**. Anti-amyloid immunotherapy prescription was associated with lower unadjusted rates of ED visits (RR 0.71, 95% CI 0.57-0.87) and hospitalizations (RR 0.62, 95% CI 0.45-0.84) However there was no association between anti-amyloid immunotherapy prescription and either ED visits (owRR 0.83, 95% CI 0.68-1.03) or hospitalizations (owRR 0.83, 95% CI 0.61-1.15) after overlap weighting.

### Exploratory End Points

There was no association between anti-amyloid immunotherapy prescription and myocarditis/pericarditis, severe skin reactions, thyroid dysfunction, hyperosmolar hyperglycemic state/diabetic ketoacidosis, or acute liver injury, although estimates were imprecise (**Table 3).** After overlap weighting, there was a higher rate of respiratory tract infections among patients prescribed anti-amyloid immunotherapy (owHR 1.57 [95% CI 1.04-2.37]; adjusted rate 85.1 1,000 p-y vs. 53.6 per 1,000 p-y, adjusted rate difference 31.5 per 1,000 p-y, number needed to harm (NNH) 32).

**Table 3.** Exploratory and falsification endpoints for the study.

|  | **AChEI/Memantine** |  |  | **Amyloid Immunotherapy** |  |  |  |
| --- | --- | --- | --- | --- | --- | --- | --- |
| Outcome | Number of events | Number of person-years | Event rate | Number of events | Number of person-years | Event rate | Hazard Ratio (95% CI) |
| **Respiratory Infection** |  |  |  |  |  |  |  |
| Crude | 11060 | 204563 | 54.07 | 23 | 269 | 85.60 | 1.58 (1.05–2.37) |
| Overlap-weighted |  |  | 53.64 |  |  | 85.05 | 1.57 (1.04–2.37) |
| **Thyroid disorders** |  |  |  |  |  |  |  |
| Crude | 1798 | 201669 | 8.92 | <11 | <1537 | 7.16 | 0.80 (0.20–3.20) |
| Overlap-weighted |  |  | 7.81 |  |  | 7.12 | 0.90 (0.22–3.59) |
| **Interstitial lung disease** |  |  |  |  |  |  |  |
| Crude | 60 | 245129 | 0.24 | 0 | <280 | 0 | NA |
| Overlap-weighted |  |  | 0.14 |  |  | 0 | NA |
| **Severe skin disorders** |  |  |  |  |  |  |  |
| Crude | 252 | 241880 | 1.04 | 0 | <280 | 0 | NA |
| Overlap-weighted |  |  | 1.05 |  |  | 0 | NA |
| **Acute liver injury** |  |  |  |  |  |  |  |
| Crude | 108 | 244722 | 0.44 | <11 | <280 | 0 | NA |
| Overlap-weighted |  |  | 0.23 |  |  | 0 | NA |
| **Hyperosmolar Hyperglycemic State or DKA** |  |  |  |  |  |  |  |
| Crude | 395 | 243148 | 1.62 | 0 | <280 | 0 | NA |
| Overlap-weighted |  |  | 0 |  |  | 0 | NA |
| **Myocarditis or Pericarditis** |  |  |  |  |  |  |  |
| Crude | 23 | 245360 | 0.09 | 0 | <280 | 0 | NA |
| Overlap-weighted |  |  | 0.11 |  |  | 0 | NA |
| **Falsification Endpoint** |  |  |  |  |  |  |  |
| **Cataract** |  |  |  |  |  |  |  |
| Crude | 1838 | 229664 | 8.00 | <11 | 3506 | 3.14 | 0.39 (0.05–2.77) |
| Overlap-weighted |  |  | 7.94 |  |  | 3.04 | 0.42 (0.06–2.94) |

### Quantitative Bias Analysis/Sensitivity Analyses

The E-value for the point estimate of the primary end point to reveal a true hazard ratio of 2.0 was 4.92, suggesting that results were robust to unmeasured confounding.^24^ The E-value for the point estimate for the headache endpoint was 3.99, and the E-value for the respiratory tract infection point estimate was 2.52. The falsification end point of cataract revealed a similar rate of cataract among patients in the two groups before and after overlap weighting (**Figure 3).**

Sensitivity analyses using a clone-censor-weight approach to account for treatment switching are shown in **Supplemental Table 4**, and results were consistent with the primary analysis, although the associations with headache and respiratory tract infections were not observed in this analysis.^25^ A sensitivity analysis restricting to patients with age <75 is shown in **Supplemental Table 5** and was similar to the primary analysis.

## DISCUSSION

In this nationwide cohort study of 2,616 patients who received anti-amyloid immunotherapy from July 1, 2023 to January 1, 2025, we observed significant demographic differences in patient adoption. Compared with patients receiving standard oral dementia medications (acetylcholinesterase inhibitors/memantine alone), those prescribed anti-amyloid immunotherapy were healthier at baseline and more likely to be male, white, and from socioeconomically advantaged areas. This study did not find an association between anti-amyloid immunotherapy and increased rates of intracranial hemorrhage, ischemic stroke, or myocardial infarction. These estimates were imprecise, but they are consistent with clinical trial findings suggesting that most amyloid-related imaging abnormalities (ARIA) are asymptomatic or present with minor symptoms rather than catastrophic bleeding events. The safety analysis did, however, find a two-fold increased risk of headaches (a known adverse effect of anti-amyloid immunotherapy) and a modestly increased risk of respiratory infection, both of which merit further study.

Our findings highlight an important disparity in the real-world use of anti-amyloid immunotherapy in the United States. Prior work indicates that eligibility criteria based on amyloid biomarkers may disproportionately exclude certain racial and ethnic groups, such as Hispanic and Black patients.^26^ This is further supported by our finding of an underrepresentation of female, Hispanic, and Black patients in the immunotherapy cohort, and is consistent with findings from the AHEAD preclinical AD program demonstrating lower plasma amyloid eligibility in underrepresented racial and ethnic groups.^27^ Some evidence suggests that amyloid-beta pathology and treatment effect of anti-amyloid immunotherapy may vary by sex, and that female patients may have earlier tau deposition than male patients.^28–30^ Additionally, access to medical care, acceptability of therapy, timeliness of diagnosis, or other social and environmental factors could impact therapy adoption.^11,14,31–35^ Given the low observed prescription of anti-amyloid immunotherapy among minoritized patient populations, this study is limited in its ability to evaluate distinct safety signals in these groups.

The safety profile observed in this study is largely reassuring. While this study was unable to evaluate the rates of ARIA-H or ARIA-E, the finding of no increased rates of intracranial hemorrhage or ischemic stroke aligns with clinical trial data, suggesting that most ARIA events are asymptomatic or cause only minor symptoms rather than catastrophic bleeding.^36–38^ This study excluded a number of patients at high risk for ARIA based on established factors, such as antithrombotic use.^37,39^ Therefore, our findings should be interpreted with caution, as many patients with clear indications for anticoagulation or a history of anticoagulant therapy were excluded from the immunotherapy group. Our finding of a higher rate of headache is consistent with prior clinical trials of amyloid immunotherapies. Because headache is a common symptom of ARIA, our finding may serve as a surrogate marker for these events.^36,39^

Our study did not find an increased risk of diarrheal illness, which is a known adverse effect of anti-amyloid immunotherapy. This finding is likely due to our use of acetylcholinesterase inhibitors - which are classically associated with diarrhea - as the active control.^40^ Thus, our study establishes that anti-amyloid therapies are not associated with increased risk of diarrheal illness compared to a common alternative treatment. Additionally, we investigated a broad range of immune-related safety signals. Aside from respiratory tract infections, we found no new safety signals associated with anti-amyloid immunotherapy. The higher rates of respiratory infections observed in our study are consistent with some clinical trials, although these trials may be difficult to interpret given the general prevalence of such infections and the timing of some trials during the COVID-19 pandemic.^2,4^ Given that immunomodulatory therapies are consistently associated with elevated risk of respiratory symptoms,^41^ these findings suggest that the risk of respiratory tract infections after anti-amyloid immunotherapy warrants further investigation.

This study has several limitations. First, identifying a suitable active comparator for anti-amyloid immunotherapy is challenging. Acetylcholinesterase inhibitors/memantine are used in a wider range of patient populations, including patients without Alzheimer’s Disease and those with more advanced cognitive impairment than patients eligible for amyloid immunotherapies. Since cognitive measures are generally unavailable in semi-structured electronic health records, our study could not control for cognitive stage at entry and could accordingly not estimate the comparative effectiveness of amyloid immunotherapies. Future studies using natural language processing tools to extract cognitive testing results from unstructured electronic health records could help extend the impact and rigor of observational comparative research into these questions. Second, concurrent therapy use was very common, with most patients on amyloid immunotherapies also receiving AChEI/Memantine. This was generally prior to the initiation of anti-amyloid immunotherapy, and results were similar with a clone-censor-weight approach. Third, our study could not directly evaluate ARIA-H or ARIA-E, adverse effects unique to amyloid immunotherapies that are monitored by MRI in routine clinical practice. Fourth, our study only included patients seen at health systems using the Epic electronic health record system. This may limit generalizability to other clinical settings. Events occurring outside of Epic Health systems were also not available in our dataset. Fifth, despite the largest nationwide sample to date, this study was underpowered to detect small but clinically meaningful safety signals. This is due to low event rates in the anti-amyloid immunotherapy group and the short follow-up time given the recent approval of these therapies.

In conclusion, this study found significant demographic differences in the adoption of anti-amyloid immunotherapy and observed a generally reassuring safety profile, with no evidence of an association between anti-amyloid immunotherapy and intracranial hemorrhage or ischemic stroke, although estimates were imprecise.

## Supporting information

RECORD Checklist

TARGET Checklist

Statistical Analysis Plan

## Notes

**Funding Sources and Role of the Sponsor:** This work was funded by the Duke-UNC Alzheimer’s Disease Research Center via a Research Education Component grant to Dr. Lusk, (NIA P30AG072958) and by the Duke University Office of the Provost. The content is solely the responsibility of the authors and does not necessarily represent the official views of the National Institutes of Health. The funding agency had no role in the study design, data collection and analysis, decision to publish, or preparation of the manuscript.

### Competing Interest Statement

JBL discloses grant funding from the Alzheimer's Association, National Institute on Aging, American Heart Association, and the Duke University Office of the Provost and committee service (uncompensated) for the American Heart Association.
KVF reports no relevant disclosures.
KGJ discloses research consulting for the University of Southern California, speaker fees from Eisai Inc., and primary investigator relationships with Eisai Inc, LEXEO Therapeutics, Athira Pharma, Annovis Bio, and the Critical Path Institute.
AL discloses consulting for Lucent.
JLL reports no relevant disclosures.
LET reports no relevant disclosures.
RM discloses grant funding from Duke University Office of the Provost, Arnold Ventures, Washington Center for Equitable Growth, and the American Investment Council. McDevitt has received consulting fees through the American Society of Nephrology and Charles River Associates as well as speaker fees from Welsh, Carson, Anderson, & Stowe, InTandem Capital, and Heritage Group.
AZ reports no relevant disclosures.
HW reports no relevant disclosures.
RO reports no relevant disclosures.
SA reports no relevant disclosures.
BGH reports no relevant disclosures.
BM discloses research funding from grant K23HL161426 from the National Heart, Lung, and Blood Institute, grant 23MRFSCD1077188 from the American Heart Association, grant 2835124 from the Duke Office of Physician Scientist Development, and through Bayer Pharmaceuticals as a site principal investigator.
FL reports no relevant disclosures.
EO discloses research funding from the Alzheimer's Association.

### Funding Statement

This work was funded by the Duke-UNC Alzheimer's Disease Research Center via a Research Education Component grant to Dr. Lusk (NIA P30AG072958) and by the Duke University Office of the Provost. The content is solely the responsibility of the authors and does not necessarily represent the official views of the National Institutes of Health.

### Author Declarations

The Institutional Review Board of the University of North Carolina Chapel Hill waived ethical approval for this work (review number 459493).

