## Supplemental Tables for "Uptake and Safety Profile of Anti-Amyloid Immunotherapies in Routine Clinical Practice"

**Supplemental Table 1. Characteristics of patients who were only prescribed anti-amyloid immunotherapy versus those prescribed only acetylcholinesterase inhibitors or memantine.**

| **Variable** | **AChEI/**  **Memantine alone (n=1,065,192)** | **Anti-Amyloid Immunotherapy (n=603)** | **SMD** |
| --- | --- | --- | --- |
| **Demographics** | | | |
| Age | 78.55 | 74.24 | 0.466 |
| Sex | | | |
| Female, n (%) | 616811 (57.9%) | 303 (50.2%) | 0.154 |
| Male, n (%) | 448271 (42.1%) | 300 (49.8%) | 0.154 |
| Race | | | |
| American Indian or Alaska Native, n (%) | 2406 (0.2%) | <11 (0%) | 0.067 |
| Asian, n (%) | 22406 (2.1%) | <11 (0%) | 0.074 |
| Black or African American, n (%) | 105482 (9.9%) | <11 (0%) | 0.390 |
| More than one Race, n (%) | 96274 (9.0%) | 65 (10.8%) | 0.058 |
| Native Hawaiian or Other Pacific Islander, n (%) | 887 (0.1%) | <11 (0%) | 0.023 |
| Other Race, n (%) | 26852 (2.5%) | <11 (0%) | 0.132 |
| White, n (%) | 789622 (74.1%) | 512 (84.9%) | 0.270 |
| Ethnicity | | | |
| Hispanic or Latino, n (%) | 71081 (6.7%) | 14 (2.3%) | 0.217 |
| Not Hispanic or Latino, n (%) | 942387 (88.5%) | 560 (92.9%) | 0.217 |
| Marital status | | | |
| Divorced or Legally Separated, n (%) | 98631 (9.3%) | 25 (4.1%) | 0.208 |
| Married, n (%) | 541683 (50.9%) | 475 (78.8%) | 0.609 |
| Other, n (%) | 32543 (3.1%) | 15 (2.5%) | 0.036 |
| Unmarried, n (%) | 126863 (11.9%) | 37 (6.1%) | 0.205 |
| Widowed, n (%) | 256245 (24.1%) | 49 (8.1%) | 0.448 |
| Rural-urban commuting area (RUCA) group | | | |
| Rural, n (%) | 50929 (4.8%) | 11 (1.8%) | 0.168 |
| Suburban, n (%) | 99634 (9.4%) | 39 (6.5%) | 0.109 |
| Urban, n (%) | 896642 (84.2%) | 546 (90.5%) | 0.189 |
| U.S. Census division | | | |
| East North Central, n (%) | 204904 (19.2%) | 75 (12.4%) | 0.189 |
| East South Central, n (%) | 70929 (6.7%) | 36 (6.0%) | 0.029 |
| Middle Atlantic, n (%) | 132402 (12.4%) | 75 (12.4%) | 0.001 |
| Mountain, n (%) | 49791 (4.7%) | 25 (4.1%) | 0.026 |
| New England, n (%) | 54687 (5.1%) | 40 (6.6%) | 0.063 |
| Pacific, n (%) | 72721 (6.8%) | 31 (5.1%) | 0.072 |
| South Atlantic, n (%) | 268872 (25.2%) | 181 (30.0%) | 0.106 |
| West North Central, n (%) | 63648 (6.0%) | 31 (5.1%) | 0.037 |
| West South Central, n (%) | 135417 (12.7%) | 104 (17.2%) | 0.127 |
| Social Vulnerability Index (percentile, 2020, by ZIP) | 0.56 | 0.45 | 0.421 |
| **Laboratory variables** | | | |
| High-density lipoprotein, mg/dL | 54.56 | 59.37 | 0.268 |
| Low-density lipoprotein, mg/dL | 87.72 | 90.86 | 0.081 |
| Serum Triglycerides, mg/dL | 122.87 | 114.73 | 0.108 |
| Hematocrit (%) | 38.86 | 41.40 | 0.524 |
| Hemoglobin, mg/dL | 12.73 | 13.67 | 0.56 |
| White blood cell count, cells/ul | 8.12 | 7.19 | 0.274 |
| Platelet count, cells/ul | 232.52 | 233.67 | 0.015 |
| Potassium, mEq/L | 4.18 | 4.30 | 0.26 |
| Sodium, mEq/L | 139.25 | 139.29 | 0.014 |
| Serum Creatinine, mg/dL | 1.14 | 1.00 | 0.221 |
| ALT, u/L | 22.59 | 22.42 | 0.006 |
| AST, u/L | 27.33 | 24.41 | 0.096 |
| HbA1c (%) | 6.46 | 6.25 | 0.169 |
| **Medical history** | | | |
| Myocardial infarction, n (%) | 62516 (5.9%) | 16 (2.7%) | 0.16 |
| Congestive heart failure, n (%) | 137393 (12.9%) | 32 (5.3%) | 0.266 |
| Peripheral vascular disease, n (%) | 116374 (10.9%) | 41 (6.8%) | 0.146 |
| Cerebrovascular disease, n (%) | 165915 (15.6%) | 81 (13.4%) | 0.061 |
| Chronic obstructive pulmonary disease (COPD), n (%) | 158510 (14.9%) | 36 (6.0%) | 0.295 |
| Rheumatologic disease, n (%) | 31478 (3.0%) | 15 (2.5%) | 0.029 |
| Peptic ulcer disease, n (%) | 9820 (0.9%) | <11 (0%) | 0.075 |
| Mild liver disease, n (%) | 35398 (3.3%) | 16 (2.7%) | 0.039 |
| Diabetes mellitus (no complications), n (%) | 224053 (21.0%) | 75 (12.4%) | 0.232 |
| Diabetes mellitus with complications, n (%) | 138267 (13.0%) | 43 (7.1%) | 0.195 |
| Paralysis, n (%) | 16615 (1.6%) | <11 (0%) | 0.086 |
| Chronic kidney disease, n (%) | 197198 (18.5%) | 50 (8.3%) | 0.303 |
| Any malignancy (excluding skin), n (%) | 95354 (9.0%) | 45 (7.5%) | 0.054 |
| Moderate to severe liver disease, n (%) | 3417 (0.3%) | <11 (0%) | 0.031 |
| Metastatic solid tumor, n (%) | 19584 (1.8%) | <11 (0%) | 0.146 |
| HIV/AIDS, n (%) | 1395 (0.1%) | <11 (0%) | 0.009 |
| Cardiac arrhythmia, n (%) | 285298 (26.8%) | 92 (15.3%) | 0.286 |
| Depression, n (%) | 230530 (21.6%) | 101 (16.7%) | 0.124 |
| Drug use, n (%) | 19686 (1.8%) | <11 (0%) | 0.126 |
| Psychoses, n (%) | 35725 (3.4%) | 23 (3.8%) | 0.025 |
| Deep venous thrombosis, n (%) | 4537 (0.4%) | <11 (0%) | 0.092 |
| Valvular heart disease, n (%) | 103159 (9.7%) | 50 (8.3%) | 0.049 |
| Atrial fibrillation, n (%) | 158828 (14.9%) | 47 (7.8%) | 0.226 |
| Atrial flutter, n (%) | 17501 (1.6%) | <11 (0%) | 0.111 |
| Alcohol use disorder, n (%) | 16430 (1.5%) | <11 (0%) | 0.033 |
| Anxiety disorders, n (%) | 192159 (18.0%) | 91 (15.1%) | 0.079 |
| Chronic pain, n (%) | 286131 (26.9%) | 148 (24.5%) | 0.053 |
| Epilepsy, n (%) | 36548 (3.4%) | <11 (0%) | 0.196 |
| Fall-related injury, n (%) | 200038 (18.8%) | 74 (12.3%) | 0.180 |
| Malnutrition, n (%) | 92038 (8.6%) | 24 (4.0%) | 0.193 |
| Migraine, n (%) | 28430 (2.7%) | 17 (2.8%) | 0.009 |
| Osteoarthritis, n (%) | 154399 (14.5%) | 73 (12.1%) | 0.070 |
| Peripheral artery disease, n (%) | 111164 (10.4%) | 35 (5.8%) | 0.170 |
| Pneumonia, n (%) | 55140 (5.2%) | <11 (0%) | 0.27 |
| Urinary tract infection, n (%) | 113911 (10.7%) | 21 (3.5%) | 0.284 |
| **Procedures** | | | |
| Coronary artery bypass grafting (CABG), n (%) | 434 (0.0%) | <11 (0%) | 0.033 |
| Electrical cardioversion, n (%) | 2066 (0.2%) | <11 (0%) | 0.017 |
| Pacemaker, n (%) | 16563 (1.6%) | <11 (0%) | 0.034 |
| Defibrillator, n (%) | 4364 (0.4%) | <11 (0%) | 0.028 |
| Implantable cardiac monitor, n (%) | 4104 (0.4%) | <11 (0%) | 0.065 |
| Cardiac valve surgery, n (%) | 1214 (0.1%) | <11 (0%) | 0.056 |
| Percutaneous coronary intervention (stent), n (%) | 3292 (0.3%) | <11 (0%) | 0.019 |
| Peripheral arterial stent, n (%) | 1411 (0.1%) | <11 (0%) | 0.001 |
| **Prescription drug exposures** | | | |
| Angiotensin receptor blocker (ARB), n (%) | 226061 (21.2%) | 137 (22.7%) | 0.036 |
| ACE inhibitor, n (%) | 200477 (18.8%) | 84 (13.9%) | 0.132 |
| Calcium channel blocker, n (%) | 293988 (27.6%) | 109 (18.1%) | 0.228 |
| Beta-blocker, n (%) | 388118 (36.4%) | 165 (27.4%) | 0.196 |
| Potassium-sparing diuretic, n (%) | 51517 (4.8%) | 25 (4.1%) | 0.033 |
| Thiazide diuretic, n (%) | 134339 (12.6%) | 51 (8.5%) | 0.136 |
| Loop diuretic, n (%) | 175578 (16.5%) | 42 (7.0%) | 0.299 |
| Alpha-1 antagonist, n (%) | 21559 (2.0%) | 13 (2.2%) | 0.009 |
| Vasodilator, n (%) | 154544 (14.5%) | 52 (8.6%) | 0.185 |
| Anticoagulant, n (%) | 160447 (15.1%) | 36 (6.0%) | 0.3 |
| Antiplatelets, n (%) | 288794 (27.1%) | 130 (21.6%) | 0.13 |
| Statin, n (%) | 559015 (52.5%) | 314 (52.1%) | 0.008 |
| Fibrates, n (%) | 20629 (1.9%) | 11 (1.8%) | 0.008 |
| Bile acid sequestrant, n (%) | 15342 (1.4%) | <11 (0%) | 0.004 |
| PCSK9 inhibitor, n (%) | 6502 (0.6%) | 26 (4.3%) | 0.241 |
| Ezetimibe, n (%) | 38715 (3.6%) | 44 (7.3%) | 0.162 |
| Icosapent ethyl, n (%) | 5463 (0.5%) | <11 (0%) | 0.056 |
| Metformin, n (%) | 123337 (11.6%) | 63 (10.4%) | 0.036 |
| Insulin, n (%) | 160801 (15.1%) | 50 (8.3%) | 0.213 |
| GLP-1 receptor Agonists, n (%) | 41037 (3.9%) | 25 (4.1%) | 0.015 |
| SGLT-2 Inhibitors, n (%) | 52819 (5.0%) | 32 (5.3%) | 0.016 |
| Sulfonylureas, n (%) | 49617 (4.7%) | 13 (2.2%) | 0.138 |
| DDP-IV Inhibitors, n (%) | 31281 (2.9%) | 11 (1.8%) | 0.073 |
| Thiazolidinediones, n (%) | 11733 (1.1%) | <11 (0%) | 0.006 |
| SSRI, n (%) | 300844 (28.2%) | 172 (28.5%) | 0.006 |
| SNRI, n (%) | 103454 (9.7%) | 62 (10.3%) | 0.019 |
| Tricyclic antidepressant (TCA), n (%) | 37893 (3.6%) | 14 (2.3%) | 0.073 |
| Monoamine oxidase inhibitor, n (%) | 1713 (0.2%) | <11 (0%) | 0.057 |
| Buspirone, n (%) | 47476 (4.5%) | 38 (6.3%) | 0.082 |
| Benzodiazepines, n (%) | 295224 (27.7%) | 150 (24.9%) | 0.065 |
| Trazodone, n (%) | 122810 (11.5%) | 52 (8.6%) | 0.097 |
| Bupropion, n (%) | 54877 (5.2%) | 47 (7.8%) | 0.108 |
| Mirtazapine, n (%) | 76145 (7.1%) | <11 (0%) | 0.303 |
| Gaba Analogs, n (%) | 197340 (18.5%) | 77 (12.8%) | 0.159 |
| Typical Antipsychotics, n (%) | 50468 (4.7%) | <11 (0%) | 0.176 |
| Second-generation antipsychotic, n (%) | 168863 (15.9%) | 14 (2.3%) | 0.484 |
| Thyroid hormone replacement, n (%) | 181989 (17.1%) | 82 (13.6%) | 0.097 |
| Bisphosphonate, n (%) | 52874 (5.0%) | 28 (4.6%) | 0.015 |
| Short-acting β-agonist (SABA), n (%) | 231652 (21.7%) | 75 (12.4%) | 0.249 |
| Short-acting muscarinic antagonist (SAMA), n (%) | 127479 (12.0%) | 34 (5.6%) | 0.225 |
| Long-acting β-agonist (LABA), n (%) | 78594 (7.4%) | 29 (4.8%) | 0.108 |
| Long-acting muscarinic antagonist (LAMA), n (%) | 82496 (7.7%) | 28 (4.6%) | 0.129 |
| Inhaled corticosteroid, n (%) | 158163 (14.8%) | 91 (15.1%) | 0.007 |
| Proton pump inhibitor, n (%) | 318146 (29.9%) | 139 (23.1%) | 0.155 |
| H2 receptor blocker, n (%) | 144946 (13.6%) | 69 (11.4%) | 0.065 |
| Opioids, n (%) | 408407 (38.3%) | 194 (32.2%) | 0.129 |
| Penicillin (± beta-lactamase inhibitor), n (%) | 140611 (13.2%) | 80 (13.3%) | 0.002 |
| First Generation Cephalosporins, n (%) | 194752 (18.3%) | 75 (12.4%) | 0.163 |
| 2nd-generation cephalosporin, n (%) | 28700 (2.7%) | 15 (2.5%) | 0.013 |
| 3rd-generation cephalosporin, n (%) | 96408 (9.1%) | 32 (5.3%) | 0.145 |
| Fourth Generation Cephalosporins, n (%) | 12403 (1.2%) | <11 (0%) | 0.123 |
| Fifth Generation Cephalosporins, n (%) | 22 (0.0%) | <11 (0%) | 0.006 |
| Carbapenem, n (%) | 3909 (0.4%) | <11 (0%) | 0.039 |
| Monobactam, n (%) | 412 (0.0%) | <11 (0%) | 0.028 |
| Macrolide, n (%) | 96750 (9.1%) | 53 (8.8%) | 0.01 |
| Fluoroquinolones, n (%) | 129300 (12.1%) | 58 (9.6%) | 0.081 |
| Tetracycline, n (%) | 106518 (10.0%) | 66 (10.9%) | 0.031 |
| Aminoglycoside, n (%) | 19047 (1.8%) | 15 (2.5%) | 0.048 |
| Sulfonamide, n (%) | 6918 (0.6%) | <11 (0%) | 0.02 |
| Lincosamide, n (%) | 32640 (3.1%) | 22 (3.6%) | 0.032 |
| Glycopeptide, n (%) | 52285 (4.9%) | 13 (2.2%) | 0.15 |
| Oxazolidinone, n (%) | 5570 (0.5%) | <11 (0%) | 0.029 |
| Nitroimidazole, n (%) | 44536 (4.2%) | 23 (3.8%) | 0.019 |
| **Healthcare utilization** | | | |
| Hospital admissions | 0.3 | 0.07 | 0.458 |
| Emergency department visits | 0.62 | 0.22 | 0.417 |
| Outpatient face-to-face visits | 4.91 | 7.73 | 0.321 |
| Primary Care visits | 1.15 | 1.03 | 0.053 |
| Neurology visits | 0.4 | 1.01 | 0.459 |
| Psychiatry visits | 0.09 | 0.11 | 0.025 |
| Geriatrics visits | 0.09 | 0.06 | 0.016 |
| Cardiology visits | 0.41 | 0.71 | 0.142 |
| Hospice/palliative care visits | 0.03 | 0 | 0.026 |
| **Vitals** | | | |
| Body Mass Index (kg/m²), median value | 27.18 | 26.64 | 0.098 |
| Diastolic blood pressure (mmHg), median value | 72.79 | 73.1 | 0.036 |
| Pulse rate (beats/min), median value | 73.29 | 70.16 | 0.276 |
| Oxygen saturation (SpO₂, %), median value | 96.8 | 97.3 | 0.299 |
| Systolic blood pressure (mmHg), median value | 132.34 | 129.25 | 0.197 |
| Body Mass Index (kg/m²), closest-to-index value | 27.1 | 26.61 | 0.089 |
| Calculated Body Mass Index (kg/m²), closest-to-index value | 27.07 | 26.59 | 0.087 |
| Diastolic blood pressure (mmHg), closest-to-index value | 72.81 | 72.69 | 0.011 |
| Pulse rate (beats/min), closest-to-index value | 73.52 | 71.25 | 0.172 |
| Oxygen saturation (SpO₂, %), closest-to-index value | 96.72 | 97.24 | 0.252 |
| Systolic blood pressure (mmHg), closest-to-index value | 131.32 | 128.26 | 0.164 |
| Weight (lbs), closest-to-index value | 166.14 | 169.52 | 0.085 |
| Respiratory rate (breaths/min), closest-to-index value | 17.13 | 16.97 | 0.075 |
| Temperature (°F), closest-to-index value | 97.79 | 97.82 | 0.042 |
| Systolic blood pressure (mmHg), median ≥130, n (%) | 352291 (33.1%) | 251 (41.6%) | 0.192 |
| Systolic blood pressure (mmHg), median ≥140, n (%) | 194209 (18.2%) | 115 (19.1%) | 0.218 |
| Systolic blood pressure (mmHg), max ≥160, n (%) | 216238 (20.3%) | 110 (18.2%) | 0.313 |
| Systolic blood pressure (mmHg), max ≥180, n (%) | 90257 (8.5%) | 30 (5.0%) | 0.292 |
| Systolic blood pressure (mmHg), min ≤89, n (%) | 38126 (3.6%) | <11 (0%) | 0.237 |
| Oxygen saturation (SpO₂, %), min ≤91, n (%) | 90064 (8.5%) | 24 (4.0%) | 0.35 |
| Oxygen saturation (SpO₂, %), min ≤89, n (%) | 40882 (3.8%) | <11 (0%) | 0.278 |
| Diastolic blood pressure (mmHg), median ≥80, n (%) | 147754 (13.9%) | 127 (21.1%) | 0.005 |
| Diastolic blood pressure (mmHg), median ≥90, n (%) | 23213 (2.2%) | 13 (2.2%) | 0.074 |
| Diastolic blood pressure (mmHg), max ≥100, n (%) | 81356 (7.6%) | 23 (3.8%) | 0.307 |
| Diastolic blood pressure (mmHg), min ≤59, n (%) | 216757 (20.3%) | 146 (24.2%) | 0.162 |

**Supplemental Table 2. Characteristics of patients who were ever prescribed anti-amyloid immunotherapy versus those prescribed only acetylcholinesterase inhibits or memantine, after propensity score overlap weighting.**

| **Variable** | **AChEI/**  **Memantine alone** | **Anti-Amyloid Immuno-therapy** | **SMD** |
| --- | --- | --- | --- |
| **Demographics** | | | |
| Age | 73.51 | 73.51 | 0 |
| Sex | | | |
| Female, n (%) | 55.7% | 55.7% | 0 |
| Male, n (%) | 44.3% | 44.3% | 0 |
| Race | | | |
| American Indian or Alaska Native, n (%) | 0.0% | 0.0% | 0 |
| Asian, n (%) | 1.0% | 1.0% | 0 |
| Black or African American, n (%) | 1.8% | 1.8% | 0 |
| More than one Race, n (%) | 11.2% | 11.2% | 0 |
| Native Hawaiian or Other Pacific Islander, n (%) | 0.2% | 0.2% | 0 |
| Other Race, n (%) | 0.8% | 0.8% | 0 |
| White, n (%) | 84.5% | 84.5% | 0 |
| Ethnicity | | | |
| Hispanic or Latino, n (%) | 1.5% | 1.5% | 0 |
| Not Hispanic or Latino, n (%) | 98.5% | 98.5% | 0 |
| Marital status | | | |
| Divorced or Legally Separated, n (%) | 4.3% | 4.3% | 0 |
| Married, n (%) | 76.9% | 76.9% | 0 |
| Other, n (%) | 2.0% | 2.0% | 0 |
| Unmarried, n (%) | 7.8% | 7.8% | 0 |
| Widowed, n (%) | 9.0% | 9.0% | 0 |
| Rural-urban commuting area (RUCA) group | | | |
| Rural, n (%) | 1.8% | 1.8% | 0 |
| Suburban, n (%) | 6.7% | 6.7% | 0 |
| Urban, n (%) | 91.5% | 91.5% | 0 |
| U.S. Census division | | | |
| East North Central, n (%) | 13.0% | 13.0% | 0 |
| East South Central, n (%) | 5.5% | 5.5% | 0 |
| Middle Atlantic, n (%) | 15.7% | 15.7% | 0 |
| Mountain, n (%) | 3.0% | 3.0% | 0 |
| New England, n (%) | 7.6% | 7.6% | 0 |
| Pacific, n (%) | 5.2% | 5.2% | 0 |
| South Atlantic, n (%) | 30.5% | 30.5% | 0 |
| West North Central, n (%) | 4.3% | 4.3% | 0 |
| West South Central, n (%) | 15.2% | 15.2% | 0 |
| Social Vulnerability Index (percentile, 2020, by ZIP) | 0.45 | 0.45 | 0 |
| **Laboratory variables** | | | |
| High-density lipoprotein, mg/dL | 58.61 | 58.61 | 0 |
| Low-density lipoprotein, mg/dL | 97.72 | 97.72 | 0 |
| Serum Triglycerides, mg/dL | 113.31 | 113.31 | 0 |
| Hematocrit (%) | 42.23 | 42.23 | 0 |
| Hemoglobin, mg/dL | 13.95 | 13.95 | 0 |
| White blood cell count, cells/ul | 6.77 | 6.77 | 0 |
| Platelet count, cells/ul | 236.82 | 236.82 | 0 |
| Potassium, mEq/L | 4.29 | 4.29 | 0 |
| Sodium, mEq/L | 139.73 | 139.73 | 0 |
| Serum Creatinine, mg/dL | 0.93 | 0.93 | 0 |
| ALT, u/L | 20.36 | 20.36 | 0 |
| AST, u/L | 22.8 | 22.8 | 0 |
| HbA1c (%) | 5.8 | 5.8 | 0 |
| **Medical history** | | | |
| Congestive heart failure, n (%) | 2.0% | 2.0% | 0 |
| Peripheral vascular disease, n (%) | 7.7% | 7.7% | 0 |
| Chronic obstructive pulmonary disease (COPD), n (%) | 5.8% | 5.8% | 0 |
| Diabetes mellitus (no complications), n (%) | 12.0% | 12.0% | 0 |
| Diabetes mellitus with complications, n (%) | 6.0% | 6.0% | 0 |
| Chronic kidney disease, n (%) | 5.5% | 5.5% | 0 |
| Any malignancy (excluding skin), n (%) | 6.5% | 6.5% | 0 |
| Cardiac arrhythmia, n (%) | 8.5% | 8.5% | 0 |
| Depression, n (%) | 16.0% | 16.0% | 0 |
| Valvular heart disease, n (%) | 6.2% | 6.2% | 0 |
| Anxiety disorders, n (%) | 18.0% | 18.0% | 0 |
| Chronic pain, n (%) | 25.1% | 25.1% | 0 |
| Fall-related injury, n (%) | 11.7% | 11.7% | 0 |
| Malnutrition, n (%) | 3.7% | 3.7% | 0 |
| Osteoarthritis, n (%) | 12.0% | 12.0% | 0 |
| Peripheral artery disease, n (%) | 6.0% | 6.0% | 0 |
| Urinary tract infection, n (%) | 4.5% | 4.5% | 0 |
| **Prescription drug exposures** | | | |
| Angiotensin receptor blocker (ARB), n (%) | 20.1% | 20.1% | 0 |
| ACE inhibitor, n (%) | 14.3% | 14.3% | 0 |
| Calcium channel blocker, n (%) | 16.5% | 16.5% | 0 |
| Beta-blocker, n (%) | 23.5% | 23.5% | 0 |
| Thiazide diuretic, n (%) | 9.0% | 9.0% | 0 |
| Loop diuretic, n (%) | 4.0% | 4.0% | 0 |
| Vasodilator, n (%) | 7.0% | 7.0% | 0 |
| Antiplatelets, n (%) | 17.7% | 17.7% | 0 |
| Statin, n (%) | 51.4% | 51.4% | 0 |
| Metformin, n (%) | 11.2% | 11.2% | 0 |
| Insulin, n (%) | 7.2% | 7.2% | 0 |
| SSRI, n (%) | 27.5% | 27.5% | 0 |
| SNRI, n (%) | 11.4% | 11.4% | 0 |
| Benzodiazepines, n (%) | 25.7% | 25.7% | 0 |
| Trazodone, n (%) | 8.0% | 8.0% | 0 |
| Bupropion, n (%) | 7.9% | 7.9% | 0 |
| Mirtazapine, n (%) | 2.0% | 2.0% | 0 |
| Gaba Analogs, n (%) | 13.3% | 13.3% | 0 |
| Second-generation antipsychotic, n (%) | 3.3% | 3.3% | 0 |
| Thyroid hormone replacement, n (%) | 13.5% | 13.5% | 0 |
| Bisphosphonate, n (%) | 5.0% | 5.0% | 0 |
| Short-acting β-agonist (SABA), n (%) | 12.5% | 12.5% | 0 |
| Short-acting muscarinic antagonist (SAMA), n (%) | 4.2% | 4.2% | 0 |
| Long-acting β-agonist (LABA), n (%) | 3.8% | 3.8% | 0 |
| Long-acting muscarinic antagonist (LAMA), n (%) | 3.7% | 3.7% | 0 |
| Inhaled corticosteroid, n (%) | 13.0% | 13.0% | 0 |
| Proton pump inhibitor, n (%) | 22.8% | 22.8% | 0 |
| H2 receptor blocker, n (%) | 12.9% | 12.9% | 0 |
| Opioids, n (%) | 30.4% | 30.4% | 0 |
| Penicillin (± beta-lactamase inhibitor), n (%) | 13.9% | 13.9% | 0 |
| First Generation Cephalosporins, n (%) | 11.7% | 11.7% | 0 |
| 3rd-generation cephalosporin, n (%) | 5.0% | 5.0% | 0 |
| Macrolide, n (%) | 10.4% | 10.4% | 0 |
| Fluoroquinolones, n (%) | 9.9% | 9.9% | 0 |
| Tetracycline, n (%) | 10.7% | 10.7% | 0 |
| **Healthcare utilization** | | | |
| Hospital admissions | 0.03 | 0.03 | 0 |
| Emergency department visits | 0.18 | 0.18 | 0 |
| Outpatient face-to-face visits | 7.95 | 7.95 | 0 |
| Primary Care visits | 1.18 | 1.18 | 0 |
| Neurology visits | 1.09 | 1.09 | 0 |
| Cardiology visits | 0.57 | 0.57 | 0 |
| **Vitals** | | | |
| Body Mass Index (kg/m²), median value | 26.49 | 26.49 | 0 |
| Diastolic blood pressure (mmHg), median value | 73.9 | 73.9 | 0 |
| Pulse rate (beats/min), median value | 71 | 71 | 0 |
| Oxygen saturation (SpO₂, %), median value | 97.07 | 97.07 | 0 |
| Systolic blood pressure (mmHg), median value | 129.91 | 129.91 | 0 |
| Body Mass Index (kg/m²), closest-to-index value | 26.48 | 26.48 | 0 |
| Calculated Body Mass Index (kg/m²), closest-to-index value | 26.46 | 26.46 | 0 |
| Diastolic blood pressure (mmHg), closest-to-index value | 73.41 | 73.41 | 0 |
| Pulse rate (beats/min), closest-to-index value | 71.43 | 71.43 | 0 |
| Oxygen saturation (SpO₂, %), closest-to-index value | 97.02 | 97.02 | 0 |
| Systolic blood pressure (mmHg), closest-to-index value | 129.11 | 129.11 | 0 |
| Weight (lbs), closest-to-index value | 166.8 | 166.8 | 0 |
| Respiratory rate (breaths/min), closest-to-index value | 16.77 | 16.77 | 0 |
| Temperature (°F), closest-to-index value | 97.79 | 97.79 | 0 |
| Systolic blood pressure (mmHg), median ≥130, n (%) | 45.9% | 45.9% | 0 |
| Systolic blood pressure (mmHg), median ≥140, n (%) | 20.8% | 20.8% | 0 |
| Systolic blood pressure (mmHg), max ≥160, n (%) | 18.8% | 18.8% | 0 |
| Systolic blood pressure (mmHg), max ≥180, n (%) | 5.3% | 5.3% | 0 |
| Oxygen saturation (SpO₂, %), min ≤91, n (%) | 7.5% | 7.5% | 0 |
| Diastolic blood pressure (mmHg), median ≥80, n (%) | 25.5% | 25.5% | 0 |
| Diastolic blood pressure (mmHg), max ≥100, n (%) | 3.0% | 3.0% | 0 |
| Diastolic blood pressure (mmHg), min ≤59, n (%) | 20.9% | 20.9% | 0 |

*Variables with low variance or near zero completion are excluded from the propensity score and accordingly from this table, per the statistical analysis plan.

**Supplemental Table 3. Healthcare utilization endpoints, modeled as count outcomes.**

| **Outcome** | **Rate Ratio (95% CI)** |
| --- | --- |
| **ED Visits** | |
| Crude | 0.71(0.57-0.87) |
| Overlap-weighted | 0.83 (0.68-1.03) |
| **Hospitalizations** | |
| Crude | 0.62 (0.45-0.84) |
| Overlap-weighted | 0.83 (0.61-1.15) |

**Supplemental Table 4. Sensitivity analysis. Hazard ratios for the primary, secondary, selected exploratory, and falsification study endpoints, with clone-censor weighting.**

| **Outcome** | **Hazard Ratio (95% CI)** |
| --- | --- |
| **Primary end point** |  |
| Intracranial hemorrhage | 1.00 (0.14, 7.00) |
| **Secondary end points** |  |
| Ischemic stroke | 1.70 (0.80, 3.63) |
| Myocardial infarction | NA^a^ |
| Composite of ICH, ischemic stroke, or MI | 1.19 (0.59, 2.42) |
| Diarrheal illness | 0.50 (0.22, 1.13) |
| Headache | 1.80 (0.91, 3.56) |
| First ED visit | 0.85 (0.69, 1.04) |
| First Hospitalization | 0.84 (0.62, 1.14) |
| **Exploratory end points^b^** |  |
| Respiratory infection | 1.11 (0.70, 1.78) |
| Thyroid disorders | 1.64 (0.41, 6.51) |
| **Falsification end point** |  |
| Cataract | 0.37 (0.05, 2.58) |

a. No events occurred in the treatment group, so no model was estimated
b. No events occurred in the treatment group for other exploratory endpoints, so no models were estimated

**Supplemental Table 5. Sensitivity analysis. Crude and adjusted rates and hazard ratios for the primary, secondary, exploratory, and falsification endpoints for the study, restricted to patients age <75.**

|  | **AChEI/Memantine** | | | **Amyloid Immunotherapy** | | |  |
| --- | --- | --- | --- | --- | --- | --- | --- |
| Outcome | Number of events | Number of person-years | Event rate | Number of events | Number of person-years | Event rate | Hazard Ratio (95% CI) |
| **Primary endpoint** | | | | | | | |
| **Intracranial hemorrhage** | | | | | | | |
| Crude | 933 | 244180 | 3.82 | <11 | <3732 | 2.95 | 0.77 (0.11-5.46) |
| Overlap-weighted |  |  | 4.00 |  |  | 2.97 | 0.73 (0.10-5.12) |
| **Secondary endpoints** | | | | | | | |
| **Ischemic stroke** | | | | | | | |
| Crude | 2995 | 238995 | 12.53 | <11 | <727 | 15.12 | 1.20 (0.50–2.88) |
| Overlap-weighted |  |  | 9.93 |  |  | 14.86 | 1.50 (0.62–3.68) |
| **Myocardial infarction** | | | | | | | |
| Crude | 1206 | 239771 | 5.03 | 0 | 0 | 0 | NA |
| Overlap-weighted |  |  | 3.73 |  |  | 0 | NA |
| **Composite of ICH, ischemic stroke, or MI** | | | | | | | |
| Crude | 4765 | 232668 | 20.48 | <11 | <586 | 18.76 | 0.91 (0.41-2.03) |
| Overlap-weighted |  |  | 16.61 |  |  | 18.51 | 1.11 (0.50-2.50) |
| **Diarrheal illness** | | | | | | | |
| Crude | 5689 | 222554 | 25.56 | <11 | <570 | 19.27 | 0.75 (0.34-1.67) |
| Overlap-weighted |  |  | 28.15 |  |  | 19.15 | 0.68 (0.30-1.52) |
| **Headache** | | | | | | | |
| Crude | 2909 | 211889 | 13.73 | <11 | <351 | 13.73 | 2.28 (1.18-4.38) |
| Overlap-weighted |  |  | 13.97 |  |  | 31.25 | 2.16 (1.12-4.16) |
| **Respiratory infection** | | | | | | | |
| Crude | 4528 | 80338 | 56.36 | 16 | 135 | 118.17 | 2.09 (1.28–3.41) |
| Overlap-weighted |  |  | 53.64 |  |  | 116.63 | 2.09 (1.27–3.43) |
| **Exploratory endpoints** | | | | | | | |
| **Thyroid disorders** | | | | | | | |
| Crude | 729 | 82933 | 8.79 | <11 | <789 | 13.95 | 1.58 (0.39–6.31) |
| Overlap-weighted |  |  | 6.86 |  |  | 13.86 | 1.99 (0.50–7.98) |
| **Interstitial lung disease** | | | | | | | |
| Crude | 15 | 98738 | 0.15 | <11 | <# | 0 | NA |
| Overlap-weighted |  |  | 0.08 |  |  | 0 | NA |
| **Severe skin disorders** | | | | | | | |
| Crude | 114 | 97087 | 1.17 | <11 | <# | 0 | NA |
| Overlap-weighted |  |  | 1.40 |  |  | 0 | NA |
| **Acute liver injury** | | | | | | | |
| Crude | 60 | 98355 | 0.61 | <11 | <# | 0 | NA |
| Overlap-weighted |  |  | 0.28 |  |  | 0 | NA |
| **Hyperosmolar Hyperglycemic State or DKA** | | | | | | | |
| Crude | 187 | 97754 | 1.91 | <11 | <# | 0 | NA |
| Overlap-weighted |  |  | 0.95 |  |  | 0 | NA |
| **Myocarditis or Pericarditis** | | | | | | | |
| Crude | 18 | 98757 | 0.18 | 0 | <280 | 0 | NA |
| Overlap-weighted |  |  | 0.19 |  |  | 0 | NA |
| **Falsification Endpoint** | | | | | | | |
| **Cataract** | | | | | | | |
| Crude | 745 | 92735 | 8.03 | <11 | <1807 | 6.09 | 0.75 (0.11–5.33) |
| Overlap-weighted |  |  | 7.02 |  |  | 5.92 | 0.83 (0.12–5.81) |
