## Supplementary material for "Uptake and Safety Profile of Anti-Amyloid Immunotherapies in Routine Clinical Practice": TARGET Checklist

**TARGET Checklist (adapted from Cashin et al., BMJ 2025)**

| **Item No** | **Checklist item** | | | **Location in Manuscript** |
| --- | --- | --- | --- | --- |
| **Abstract** | | | |  |
| 1 | a | Identify that the study attempts to emulate a target trial using observational data. State the study objectives and briefly summarize the specified target trial. | | 4 |
|  | b | Report the data sources used for emulation. | | 4 |
|  | c | Summarize key assumptions, statistical methods, findings and conclusions. | | 4-5 |
| **Introduction** | | | |  |
| 2 | Background | Describe the scientific background of the study and the gap in knowledge. | | 6 |
| 3 | Causal question | Summarize the causal question. | | 6 |
| 4 | Rationale | Describe the rationale for emulating a target trial with the available data. Cite randomized trials informing the design of the target trial if applicable. | | 6 |
| **Methods** | | | |  |
| 5 | Data sources | Cite the data sources contributing to the analyses and for each one describe the following: original purpose, type, the geographical locations, setting, and time period. If relevant, describe how the data were linked or pooled. | | 6 |
| **Target trial specification (item 6) and target trial emulation (item 7)** | | | |  |
| 6 | Specify the components of the target trial protocol that would answer the causal question. | 7 | Describe how the components of the target trial protocol were emulated with the observational data, including how all variables were measured or ascertained. | 6-9 |
| Eligibility criteria | | | |  |
| 6a | Describe the eligibility criteria. | 7a | Describe how the eligibility criteria were operationalized with the data. | 7 |
| Treatment strategies | | | |  |
| 6b | Describe the treatment strategies that would be compared. | 7b | Describe how the treatment strategies were operationalized with the data. | 8 |
| Assignment procedures | | | |  |
| 6c | Report that eligible individuals would be randomly assigned to treatment strategies and may be aware of their treatment allocation. | 7c | Describe how assignment to treatment strategies was operationalized with the data. | 8 |
| Follow-up | | | |  |
| 6d | Clarify that follow-up would start at time of assignment to the treatment strategies. Specify when follow-up would end. | 7d | Clarify that follow-up starts at the time individuals were assigned to the treatment strategies. Describe how the end of follow-up was operationalized with the data. | 8-10 |
| Outcomes | | | |  |
| 6e | Describe the outcomes. | 7e | Describe how the outcomes were operationalized with the data. | 8 |
| Causal contrasts | | | |  |
| 6f | Describe the causal contrasts of interest, including effect measures. | 7f | Describe how the causal contrasts were operationalized with the data, including effect measures. | 8, 10 |
| Identifying assumptions | | | |  |
| 6g | Describe assumptions that would be made to identify each causal estimand. Describe the variables, if any, related to these assumptions. | 7g.i | For each causal estimand, describe assumptions made to identify it, including assumptions regarding baseline confounding due to lack of randomization. | 10 |
|  |  | 7g.ii | Describe how the variables related to these assumptions were operationalized with the data. | 10 |
| Data analysis plan | | | |  |
| 6h | For each causal estimand, describe the data analysis procedures and any associated statistical modeling assumptions, including approaches for handling missing data. | 7h.i | For each causal estimand, describe the data analysis procedures and any associated statistical modeling assumptions, including approaches for handling missing data. | 10 |
|  |  | 7h.ii | For each causal estimand, describe any additional analyses conducted to assess the sensitivity of the results to the choice of operationalizations, assumptions and analysis. | 10 |
| **Results** | | | |  |
| 8 | Participant selection | Report numbers of individuals assessed for eligibility, eligible, and assigned to each treatment strategy. A flow diagram is strongly recommended. | | Figure 1 |
| 9 | Baseline data | Describe the distribution of characteristics of individuals at baseline, by treatment strategy. | | 11-13 |
| 10 | Follow-up | Summarize length of follow-up and describe reasons for end of follow-up for each treatment strategy and causal contrast. | | 10 |
| 11 | Missing data | Describe the frequency of missing data in all variables, by treatment strategy when applicable. | | Supplemental Material |
| 12 | Outcomes | Describe the frequency or distribution of each outcome, by treatment strategy. | | Figure 2, Table 3 |
| 13 | Effect estimates | Report the effect estimates for each causal contrast with corresponding measures of precision, including both absolute and relative measures of effect, when applicable. | | Figure 2, Table 3 |
| 14 | Additional analyses | Report results of all analyses to assess the sensitivity of the estimates to choices in operationalizations, assumptions and analysis. | | 13-14 |
| **Discussion** | | | |  |
| 15 | Interpretation | Provide an interpretation of the key findings. | | 14 |
| 16 | Limitations | Discuss the limitations of the study considering differences between the target trial and its emulation and the plausibility of assumptions, including assumptions regarding baseline confounding due to lack of randomization. | | 16-17 |
| **Other information** | | | |  |
| 17 | Ethics | Provide the institutional research board or ethics committee that approved the study and approval numbers, if relevant. | | 7 |
| 18 | Registration | State whether, when, and where the study protocol was registered. | | 6 |
| 19 | Sharing of study materials | Provide information on whether data, analytic code and/or other materials are accessible, and where and how they can be accessed. | | 7 |
| 20 | Funding sources | Provide the sources of funding and detail the role of the funders in the design, conduct and reporting of the study. | | 2 |
| 21 | Conflicts of interest | State any conflicts of interest and financial disclosures for all authors. | | 2-3 |
