## Supplementary figures and images for "Uptake and Safety Profile of Anti-Amyloid Immunotherapies in Routine Clinical Practice"

### Supplemental Figures

**eFigure 1. Crude and adjusted survival curves.**

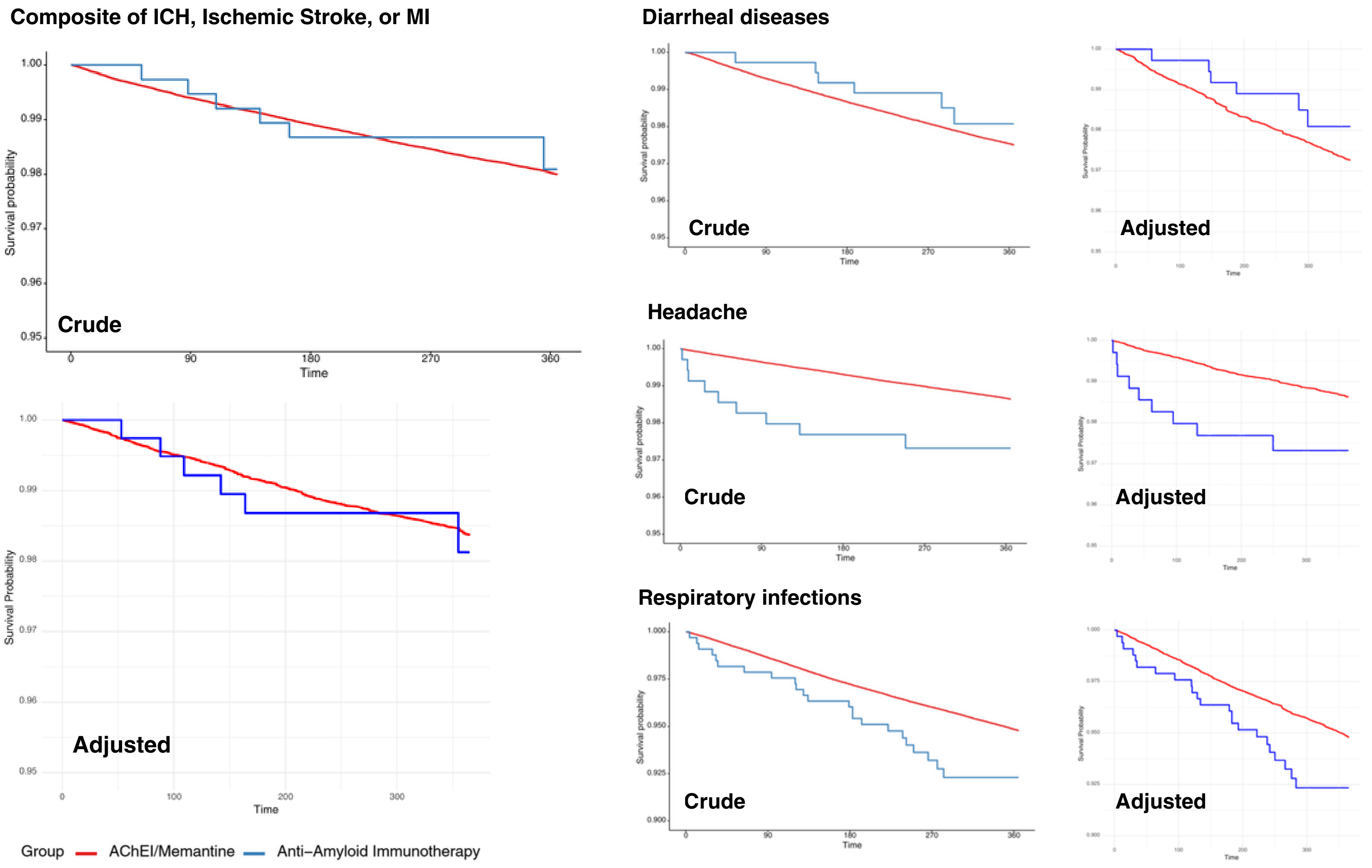
