## Supplementary material for "Uptake and Safety Profile of Anti-Amyloid Immunotherapies in Routine Clinical Practice": Statistical Analysis Plan

**Anti-Amyloid Therapy Safety Evaluation SAP**

**Objective:** To compare the frequency of adverse events after treatment initiation with anti-amyloid monoclonal antibody therapy versus approved dementia medications (donepezil, memantine, rivastigmine, or galantamine).

**Population:** Patients with dementia potentially eligible for dementia pharmacotherapy. The inclusion criterion will be first prescription of either lecanemab or donanemab (Arm 1) or donepezil, memantine, rivastigmine, or galantamine (Arm 2). The study will use a new-user active comparator design using target trial emulation principles. Accordingly, patients will be classified according to the group they are prescribed first to concord with the intent to treat principle. Patients with first prescription between 07/01/2023 and 01/01/2025 will be included.

**Exclusion Criteria:** 1) History of ischemic or hemorrhagic stroke; 2) History of atrial fibrillation; 3) Cardiac valvular surgery in lookback period; 4) Prescription for warfarin or any non-vitamin K oral anticoagulant in the lookback period; 5) Deep vein thrombosis. 6) Encounter for hospice/palliative care services in lookback period; 7) Age at index >=90 years. Codes used to identify exclusion criteria are shown in **APPENDIX 1.**

**End Points:** The primary end point will be time to first diagnosis of nontraumatic intracerebral hemorrhage, defined as I61.* or I62.* This endpoint was selected due to the known risk of amyloid-related imaging abnormalities (ARIA), which does not have a specific ICD-10 code, and is most likely to be captured as intracranial hemorrhage in electronic health record data. Codelists for each primary, secondary, and exploratory endpoint are shown in **APPENDIX 2.** For all endpoints, we will require the diagnosis code of interest in the primary diagnosis position and will not use problem list derived diagnoses. Participants with a history of each endpoint (except healthcare utilization endpoints) in the lookback window will be excluded, to avoid potential misclassification of delayed management of an incident event as a subsequent recurrent event.

*Secondary endpoints will include the following.*

1) Cardiovascular endpoints: myocardial infarction, ischemic stroke, and composite of myocardial infarction, ischemic stroke, and intracranial hemorrhage
2) Diagnosis of diarrheal illness
3) Diagnosis of headache
4) Healthcare utilization endpoints: hospitalization and emergency department visit, modeled both as a time-to-event endpoint and as a count endpoint with poisson or negative binomial regression with a 1-year offset depending on the degree of overdispersion.

The above secondary endpoints were selected as they are either 1) major preventable causes of death among the general population with dementia that could be associated with amyloid immunotherapy (i.e. cardiovascular safety endpoints); 2) Established harms of amyloid immunotherapy observed in randomized trials; 3) General healthcare utilization endpoints that reflect broader surrogate of potential harms that could manifest with a need for increased acute healthcare utilization.

*<u>Exploratory safety endpoints, based on adverse effects of other immunotherapies but not seen in existing clinical trials of amyloid immunotherapies will include the following as time-to-event endpoints</u>*

1) Lower or upper respiratory tract infection
2) Myocarditis or pericarditis
3) Acute liver injury
4) Interstitial lung disease
5) Severe skin reactions (lichen dermatitis, psoriasis, bullous pemphigoid, or SJS-TEN)
6) Hypo- or hyperthyroidism
7) Hyperosmolar hyperglycemic state or diabetic ketoacidosis

**Adjustment Covariates:**

We will use a broad range of adjustment covariates to address measured confounding, using a two-year lookback period. Specific adjustment covariates include:

1) Demographics. Age, Sex, Race, Ethnicity.
2) Social Determinants of Health. SVI percentile, ADI
3) Physical measurements. Systolic blood pressure, diastolic blood pressure, body mass index, height, weight, heart rate, oxygen saturation, and pulse. Physical measurements will be summarized as closest to index date, median in lookback, min in lookback, max in lookback, and range in lookback, to account for biologically meaningful variation in these frequently measured variables.
4) Laboratory measurements. Hemoglobin, hematocrit, white blood cell count, platelet count, aspartate aminotransferase, alanine aminotransferase, serum creatinine, sodium, potassium, glycated hemoglobin, low density lipoprotein cholesterol, high density lipoprotein cholesterol, serum triglycerides. Laboratory measurements will be selected as closest to index date within the 2-year lookback period.
5) Procedures. Coronary artery bypass grafting, carotid endarterectomy, cardiac ablation, pacemaker placement, percutaneous coronary intervention (codelist shown in **APPENDIX 3)**

6) Diagnoses. The individual components of the Elixhauser comorbidity index and claims-based frailty index, as well as history of alcohol use disorder, anxiety disorders, chronic pain, fall-related injury, epilepsy, malnutrition/unintended weight loss, migraine headache, osteoarthritis, peripheral arterial disease, pneumonia, and urinary tract infection (codelists shown in **APPENDIX 4).**
7) Common medications which either reflect general medical illness, are associated with bleeding, or are associated with cardiovascular disease (see **APPENDIX 5**).
8) Healthcare utilization. Number of outpatient encounters, emergency department encounters, hospitalizations as well as number of visits to neurologists, geriatricians, primary care physicians, and cardiologists.

Implausible laboratory measurements will be systematically excluded according to the approach detailed in **APPENDIX 6.**

**Treatment of Missing Data:** Missing data will be imputed using random forest-based imputation approach[1] using the MissRanger package, as this approach has been shown to be particularly well-suited to very high dimensional data of varying data types. We will interrogate missingness of data elements prior to performing imputation and will exclude variables with extremely high missing rates or those that are nearly universally present or absent (>99%). We will use a maximum number of tress of 100 and three candidate non-missing values to sample for predictive mean matching.

**Propensity Score Methods:** We will use propensity score overlap weighting.[1,2] If variables have near zero variance (defined systematically with the nearZeroVar function in the *caret* package in R) they will be excluded from propensity score derivation and this will be noted in a supplement.

**Statistical Analysis.** Cox proportional hazard models will be used for time to event endpoints. The maximum duration of follow-up will be set to one year. Patients will be censored at death, end of data availability, or one year after index date. Cause-specific hazards will be used, as Epic Cosmos death data is not complete making methods used to treat death as a competing risk difficult to reliably implemented.

**Quantitative Bias Analysis.**

**1)** E-values. E-values will be calculated for the primary and secondary endpoints.
**2)** Falsification endpoint. Time to diagnosis of cataract will be used as a negative control (falsification) endpoint, as cataract is a common age-related condition which requires health system engagement to address and accordingly shares underlying confounding factors with both exposures, but is not biologically linked to

either exposure. The codes used for cataract are adapted from Muir et al.[3] after conversion to ICD-10-CM, corresponding to H25.*, H26.*, H27.*, and H28.*.

**3)** Sensitivity analysis restricting to age <75 in both arms, to assess unmeasured confounding by disease severity which is most closely related to age

**4)** Sensitivity analysis using clone-censor-weighting (if feasible given number of patients in each potential treatment strategy) to emulate

  - **a.** Strategy of addition of amyloid therapy on baseline acetylcholinesterase inhibitor therapy. In this design, the index date is the date of first prescription of acetylcholinesterase therapy. Patients never receiving acetylcholinesterase inhibitors will be excluded.
  - **b.** Strategy of addition of acetylcholinesterase inhibitor therapy on baseline anti-amyloid therapy. In this design, the index date is the date of first prescription of anti-amyloid therapy. Patients never receiving anti-amyloid therapy will be excluded.
  - **c.** In both designs above, patients will be censored at the time of secondary therapy initiation in one cloned arm, and inverse probability of censoring weighting will be used to fit a weighted cox model, where the final weights are the inverse probability of censoring weights multiplied by the overlap weights.

**APPENDIX 1. Codes used to identify study exclusion criteria**

| Criterion | Codelist |
|---|---|
| History of ischemic stroke or ICH | Ischemic Stroke (Birman-Deych), G45.0, G45.1, G45.2, G45.8, G45.9, G46.0, G46.1, G46.2, I63.*, I67.81, I67.82, I67.84, I67.89, I67.9*, I69.*; ICH (Birman-Deych): @06.*, I60.*-I62.* |
| History of atrial fibrillation or atrial flutter | I48.0*-I48.2*, I48.91 or I48.3*, I48.4, I48.92 |
| Cardiac valvular surgery | CPT 33361:33478 |
| Prescription for warfarin or non-vitamin K oral anticoagulant | Generic name %like% “warfarin” or “apixaban” or “rivaroxaban” or “dabigatran” or “edoxaban” |
| Deep Vein Thrombosis | I80.1*-I80.3*, I80.9*, I82.1*, I82.210, I82.22, I82.290, I82.6*, I82.890, I82.9*, I82.A1, I82.B1, I82.C1 (Birman-Deych) |
| Hospice/Palliative Care Encounter | Department specialty of Palliative Medicine, Inpatient Hospice, Hospice Services, Home Hospice, or Hospice and Palliative Medicine |

**APPENDIX 2. Codes used to identify the primary, secondary, and exploratory endpoints.**

| Endpoint | Codelist |
|---|---|
| Intracerebral hemorrhage | I61.* or I62.* |
| Myocardial infarction | I21.* and I22.* (Metcalfe et al.[4]) |
| Ischemic stroke | I63.* (Hsieh et al.[5]) |
| Diarrheal Illness | R19.7, A09.*, K52.9*, |
| Headache | G43.*, G44.*, R51.* |
| Respiratory tract infection | J10-J18 (inclusive), Skull et al.[6]; J00-J06 |
| Myocarditis or pericarditis | A381, A3952, B2682, B3320, B3322, B3324, B5881, D8685, I012, I090, I400, I401, I408, I409, I41, I514, J1082 or J1182 (Wu et al.[7]) |
| Acute liver injury | K71.0, K71.1, K71.2, K71.6, K71.9, K72.0, K72.9, K75.9, K76.2 (Timmer et al.)[8] |
| Interstitial Lung Disease | J84.112 (Herberts et al.)[9] |
| Severe skin reactions | L51.1-L51.3 (Wasuwanich et al.[10]), L12.0 (Leisti et al.) [10], L40.0-40.9, M07.0, M07.1, M07.2, M07.3, M09.0 (Ham et al.[11]), L66.1, L43.0, L43.1, L43.2, L43.3, L43.8, L43.9 (del Toro et al)[12] |
| Hypo- or hyperthyroidism | E01.0, E01.1, E01.2, E01.8, E02.9, E03.2, E03.3, E03.8, E03.9, E06.0, E06.1, E06.2, E06.3, E06.4, E06.5, E06.9, E89.0, E89.0A, E89.0B, E89.0X (Bengtsson et al.[13]), E05.0, E05.1, E05.2, E05.8, E05.9 (Peng et al.)[14] |
| Hyperosmolar hyperglycemic state or diabetic ketoacidosis | E10.10, E11.10, E13.10, and E14.10 (DKA, Hodzic-Santor et al.[15]), E11.01, E11.00, E13.00, E08.00, E11.10 |

**APPENDIX 3: Codes used to identify procedural adjustment variables.**

| **Procedure** | **Codelist (CPT unless otherwise specified)** |
|---|---|
| Coronary artery bypass grafting | 33510, 33511, 33512, 33513, 33514, 33516, 33517, 33518, 33519, 33521, 33522, 33523, 33530, 33533, 33534, 33535, 33536 |
| Carotid endarterectomy | 33501 |
| Pacemaker | 33206, 33207, 33208, 33227, 33228, 33214, 33212, 33213, 33,229 93280, 93279, 93288, 93281 |
| Implantable defibrillator | 33249, 33240, 33230, 33231, 33262, 33263, 33264, 33270, 93289, 93282, 93283, 93284, 93641 |
| Implantable cardiac monitor | 33282, 33284, 93285, 93291, 93298 |
| Percutaneous coronary intervention | 92928, 92929, 92933, 92934, 92937, 92941, 92943, 92944; HCPCS C9600, C9601, C9602, C9603, C9604, C9605, C9606, C9607, C9608 |
| Peripheral artery stenting | CPT 37221-37223; 37226-37227; 37230-37235; 37236-37237; 37238-37239; HCPCS C1874-C1877, C2617, C2625 |

**APPENDIX 4: Codes used to identify diagnosis-based adjustment variables.**

| Condition | Codelist (ICD-10-CM) |
|---|---|
| Alcohol use disorder | F10.10, F10.11, F10.120, F10.129, F10.20, F10.21, F10.220, F10.229 |
| Anxiety disorders | F06.4*, F40.00, F40.01, F40.02, F40.10, F40.11, F40.210, F40.218, F40.220, F40.228, F40.230, F40.231, F40.232, F40.233, F40.240-F40.243, F40.248, F40.290, F40.291, F40.298, F40.8*, F40.9*, F41.0*, F41.1*, F41.3*, F41.8*, F41.9*, F42.*, F43.0*, F43.10, F43.11, F43.12, F44.9*, F45.8*, F48.8*, F48.9*, F93.8*, F99.*, R45.2*, R45.5*, R45.6*, R45.7* |
| Chronic Pain | F45.41, F45.42, , G89.0*, G89.21, G89.22, G89.28, G89.29, G89.4*, M25.50, M25.51, M25.55-M25.57, M25.78, M43.2*, M43.6*, M43.8*9, M45.9, M46.1*, M46.41-M46.47, M46.9*, M47.0*, M47.02, M47.1*, M47.24-M47.28, M47.814, M47.815, M47.816, M47.817, M47.818, M47.819, M47.894-M47.898, M48.00, M48.04-M48.08, M48.1*, M48.9*, M50.1*, M50.20, M50.3*, M50.8*, M50.9*, M51.04-M51.06, M51.1*, M51.24, M51.25, M51.2*, M51.4*, M51.8*, M51.9*, M53.1*, M53.3, M53.2*7, M53.2*8, M53.3*, M53.8*, M53.9*, M54.*, M60.8*, M60.9*, M62.830, M67.88, M79.0, M72.9*, M79.1*, M79.2*, M79.6*, M79.7*, M96.1*, M99.22-M99.29, M99.32-M99.39, M99.42-M99.49, M99.52-M99.59, M99.62-M99.69, M99.72-M99.79 |
| Epilepsy | G40.* |
| Fall-related injury | M48.4*, M48.5*, M67.90, M80.*, M84.*, M99.1*, S00.*-S99.*, T07.XXXS, T14.8XXA, T14.8XXS, T14.90XA, T14.90XS, T14.91XS, T15.00XS, T15.01XS, T15.02XS, T15.10XS, T15.11XS, T15.12XS, T15.80XS, T15.81XS,T15.82XS, T15.90XS, T15.90XS, T15.91XS, T15.92XS, T16.1XXS, T16.2XXS, T16.9XXS,T16.9XXS, T17.0XXS, T17.1XXS, T17.1XXS, T17.200S, T17.208S, T17.210S, T17.218S, T17.220S, T17.228S, T17.290S, T17.298S, T17.300S, T17.308S, T17.310S, T17.318S, T17.320S, T17.328S, T17.390S, T17.398S, T17.400S, T17.408S, T17.410S, T17.418S, T17.420S, T17.428S, T17.490S, |

| | |
|---|---|
| | T17.498S, T17.500S, T17.508S, T17.510S, T17.518S, T17.520S, T17.528S, T17.590S, T17.598S, T17.800S, T17.808S, T17.810S, T17.818S, T17.820S, T17.828S, T17.890S, T17.898S, T17.900S, T17.900S, T17.908S, T17.910S, T17.918S, T17.920S, T17.928S, T17.990S, T17.998S, T18.0XXS, T18.0XXS, T18.100S, T18.108S, T18.110S, T18.118S, T18.120S, T18.128S, T18.190S, T18.198S, T18.2XXS, T18.3XXS, T18.4XXS, T18.5XXS, T18.8XXS, T18.9XXS, T18.9XXS, T19.0XXS, T19.1XXS, T19.2XXS, T19.3XXS, T19.4XXS, T19.8XXS, T19.9XXS, T19.9XXS, T79.0XXS, T79.1XXS, T79.2XXS, T79.4XXS, T79.5XXS, T79.6XXS, T79.7XXS, T79.8XXS, T79.9XXS, T79.9XXS, T79.A0XS, T79.A11S, T79.A12S, T79.A19S, T79.A21S, T79.A22S, T79.A29S, T79.A3XS, T79.A9XS |
| Malnutrition/unintended weight loss | E40.*-E43.*, E44.0*, E44.1*, E45.*, E46.*, E64.0*, R62.0*, R62.50-R62.52, R62.59, R63.0*, R63.3*, R63.4*, R63.6*, R64.* |
| Migraine Headache | G43.* |
| Osteoarthritis | M15.*-M19.* |
| Peripheral Artery Disease | E08.52, E09.52, E10.51, E10.52, E10.59, E11.51, E11.52, E11.59, E13.51, E13.52, E13.59, I70.0*, I70.2*-I70.7*, I70.90-I70.932, I73.9*, I77.1*, I96.*, L97.*, I74.10, I74.3, I74.4, I74.5, I74.8, I75.02 |
| Pneumonia | A02.22, A20.2*, A21.2*, A22.1*, A31.0*, A37.01, A37.11, A37.81, A37.91, A42.0*, A43.0*, A48.1*, A78.*, B01.2*, B05.2*, B25.0*, B37.1*, B38.0*, B38.1*, B38.2*, B39.0*, B39.1*, B39.2*, B39.5*, B39.9*, B44.0*, B58.3*, B59.*, B77.81, J12.0*-J12.3*, J12.81, J12.89, J12.9*, J13.*-J15.*, J16.0*, J16.8*, J17.*, J18.0*, J18.1*, J18.8*, J18.9*, J85.0*, J85.1*, J85.2* |
| Urinary Tract Infection | N39.0* |

APPENDIX 5. Medications used for comorbidity adjustment.

| | |
|---|---|
| **Antihypertensives and Diuretics** | |
| Angiotensin receptor blocker (ARB) | Azilsartan, candesartan, eprosartan, irbesartan, losartan, Olmesartan, telmisartan, valsartan |
| Angiotensin-converting enzyme (ACE) inhibitor | Benazepril, captopril, enalapril, fosinopril, lisinopril, moexipril, perindopril, quinapril, ramipril, trandolapril |
| Calcium channel blocker | Amlodipine, diltiazem, felodipine, isradipine, nicardipine, nisoldipine, verapamil |
| Beta Blocker | Acebutolol, atenolol, betaxolol, bisoprolol, carvedilol, labetalol, metoprolol, nadolol, nebivolol, penbutolol, pindolol, propranolol, sotolol |
| Potassium-sparing diuretic | Triamterene, eplerenone, spironolactone, amiloride |
| Thiazide diuretic | Chlorthalidone, hydrochlorothiazide, indapamide, metolazone |
| Loop diuretic | Furosemide, bumetanide, torsemide |
| Alpha antagonists | Doxazosin, prazosin, terazosin |
| Vasodilators | Hydralazine, minoxidil |
| **Antithrombotics** | |
| Anticoagulants | Apixaban, dabigatran, edoxaban, rivaroxaban, warfarin |
| Antiplatelets | Aspirin, clopidogrel, prasugrel, ticagrelor |
| **Lipid lowering therapy** | |
| Statins | Atorvastatin, rosuvastatin, lovastatin, pitavastatin, pravastatin, simvastatin, fluvastatin |
| Fibrates | Gemfibrozil, fenofibrate, clofibrate |
| Bile acid sequestrants | Colestipol, cholestyramine resin, colesevelam |
| Proprotein convertase subtilisin/kexin type 9 (PCSK9) inhibitor | Evolocumab, Alirocumab |

| | |
|---|---|
| Ezetimibe | Ezetimibe |
| Icosapent ethyl | Omega-3 acid ethyl esters (USP) or icosapent ethyl |
| **Antiglycemics** | |
| Metformin | Metformin |
| Insulin | All insulins |
| Glucagon-like peptide (GLP)-1 agonists | Semaglutide, liraglutide, dulaglutide, exenatide, lixisenatide |
| Sodium-Glucose Cotransporter 2 (SGLT2) inhibitors | Empagliflozin, canagliflozin, ertugliflozin, dapagliflozin |
| Sulfonylureas | Glipizide, glimepiride, glyburide |
| Dispeptidyl Peptidase IV (DDP-IV) inhibitors | Sitagliptin, saxagliptin, linagliptin, alogliptin |
| Thiazolidinediones | pioglitazone, rosiglitazone |
| **Antidepressants, Anxiolytics, and Antipsychotics** | |
| SSRI | citalopram, escitalopram, fluoxetine, fluvoxamine, paroxetine, sertraline, vilazodone |
| SNRI | venlafaxine, desvenlafaxine, duloxetine, levomilnacipran, milnacipran |
| TCA | amitriptyline, amoxapine, clomipramine, desipramine, doxepin, imipramine, nortriptyline, protriptyline, trimipramine |
| MAOI | isocarboxazid, phenelzine, tranylcypromine, selegiline |
| Buspirone | Buspirone |
| Benzodiazepines | alprazolam, chlordiazepoxide, clonazepam, clorazepate, diazepam, estazolam, flurazepam, lorazepam, midazolam, oxazepam, temazepam, triazolam |
| Trazodone | Trazodone |
| Bupropion | Bupropion |
| Mirtazapine | Mirtazapine |
| GABA analogs | Pregabalin, Gabapentin |
| First-generation typical antipsychotics | fluphenazine, haloperidol, perphenazine, trifluoperazine, thiothixene, chlorpromazine, thioridazine, loxapine, molindone, mesoridazine |

| | |
|---|---|
| Second-generation antipsychotics | aripiprazole, asenapine, brexpiprazole, cariprazine, clozapine, iloperidone, lurasidone, olanzapine, paliperidone, quetiapine, risperidone, ziprasidone |
| **Endocrine** | |
| Thyroid Replacement | levothyroxine, liothyronine, liotrix, desiccated thyroid |
| Bisphosphonates | alendronate, risedronate, ibandronate, zoledronic acid, etidronate, pamidronate, tiludronate |
| **Respiratory** | |
| SABA | albuterol, levalbuterol |
| SAMA | ipratropium |
| LABA | salmeterol, formoterol, arformoterol, indacaterol, olodaterol, vilanterol |
| LAMA | tiotropium, aclidinium, umeclidinium, glycopyrrolate, revefenacin |
| ICS | beclomethasone, budesonide, ciclesonide, flunisolide, fluticasone propionate, fluticasone furoate, mometasone |
| **Gastrointestinal** | |
| Proton pump inhibitors | omeprazole, esomeprazole, lansoprazole, dexlansoprazole, pantoprazole, rabeprazole |
| Histamine receptor blockers | famotidine, cimetidine, nizatidine |
| **Opioids** | codeine, hydrocodone, oxycodone, morphine, hydromorphone, oxymorphone, fentanyl, methadone, tramadol, tapentadol, buprenorphine |
| **Antibiotics** | |
| Penicillins w/wo Beta-Lactamase inhibitors | penicillin, amoxicillin, ampicillin, oxacillin, nafcillin, dicloxacillin, piperacillin, ticarcillin, amoxicillin-clavulanate, ampicillin-sulbactam, piperacillin-tazobactam, ticarcillin-clavulanate |
| First Generation Cephalosporins | cephalexin, cefazolin |
| Second generation cephalosporins | cefuroxime, cefaclor, cefoxitin |
| Third generation cephalosporins | ceftriaxone, cefotaxime, ceftazidime, cefdinir, cefixime |
| Fourth generation cephalosporins | Cefepime |
| Fifth generation cephalosporins | ceftaroline |
| Carbapenems | imipenem, meropenem, ertapenem, doripenem |

| | |
|---|---|
| Monobactams | Aztreonam |
| Macrolides | azithromycin, clarithromycin, erythromycin |
| Fluoroquinolones | ciprofloxacin, levofloxacin, moxifloxacin, ofloxacin, delafloxacin |
| Tetracyclines | tetracycline, doxycycline, minocycline, demeclocycline, omadacycline, eravacycline |
| Aminoglycosides | gentamicin, tobramycin, amikacin, streptomycin, plazomicin |
| Sulfonamides | sulfamethoxazole-trimethoprim (SMX-TMP), sulfadiazine, sulfisoxazole |
| Lincosamides | Clindamycin |
| Glycopeptides | vancomycin, telavancin, dalbavancin, oritavancin |
| Oxazolidinones | linezolid, tedizolid |
| Nitroimidazoles | metronidazole, tinidazole |

**APPENDIX 6. Implausible measurement cutoffs.**

| Measurement | Lower limit | Upper limit |
|---|---|---|
| **Laboratory Measures** | | |
| HbA1c (%) | 3.0 | 20.0 |
| HDL Cholesterol (mg/dL) | 5 | 200 |
| LDL Cholesterol (mg/dL) | 5 | 1000 |
| Total Cholesterol (mg/dL) | 10 | 2000 |
| Serum Triglycerides (mg/dL) | 5 | 2000 |
| Serum Creatinine (mg/dL) | 0.1 | 20 |
| AST (U/L) | 1 | 2000 |
| ALT (U/L) | 1 | 2000 |
| White Blood Cell Count (cells/uL) | 0.1 | 100 |
| Hemoglobin (g/dL) | 0.5 | 30 |
| Hematocrit (%) | 10 | 90 |
| Platelet Count (thousands/uL) | 1 | 1000 |
| Potassium | 1 | 15 |
| Sodium | 100 | 255 |
| **Physical Measures** | | |
| SBP (mmHg) | 40 | 300 |
| DBP (mmHg) | 40 | 200 |
| BMI (m/kg^2) | 10 | 100 |
| Temperature (degrees Fahrenheit) | 80 | 115 |
| Pulse Rate (beats per minute) | 20 | 300 |
| Oxygen saturation (%) | 50 | 100 |
| Height (in) | 20 | 100 |
| Weight (pounds) | 40 | 1300 |
| Respiratory rate (respirations per minute) | 5 | 70 |

**DEVIATIONS (pre-modeling)**

The home oxygen, wheelchair, and home hospital bed values in the CFI, derived from procedure data, were missing at an extremely high rate, so these adjustment variables were excluded. Very few patients had preceding carotid endarterectomy or cardiac ablation, so these variables were excluded. The CFI definition of heart failure was highly collinear with the Elixhauser definition, so the CFI definition was dropped.

**ATTESTATION**

I, Jay B. Lusk, the corresponding author for this work and primary analyst, attest that this analysis will be executed faithfully according to the plan detailed above, that any deviations will be disclosed transparently in the final manuscript, and that this plan was committed prior to estimation of the primary study results. This analysis plan was locked on 07/25/2025.

Jay B. Lusk, MD

Digitally signed by Jay B. Lusk, MD
Date: 2025.07.25 12:25:10 -04'00'
